# Evaluating public health effects of risk-based travel policy for the COVID-19 epidemic in Scotland

**DOI:** 10.1101/2023.08.20.23293987

**Authors:** Isobel McLachlan, Selene Huntley, Kirstin Leslie, Jennifer Bishop, Christopher Redman, Gonzalo Yebra, Sharif Shaaban, Nicolaos Christofidis, Samantha Lycett, Matthew T.G. Holden, David L. Robertson, Alison Smith-Palmer, Joseph Hughes, Sema Nickbakhsh

**Author notes:** Corresponding authors: Dr. Sema Nickbakhsh; Dr. Alison Smith-Palmer. Shared first authors (Isobel McLachlan, Selene Huntley, Kirstin Leslie).

## Abstract

**Background:** Decisions to impose temporary travel measures are less common as the global epidemiology of COVID-19 evolves. Risk-based travel measures may avoid the need for a complete travel ban, however evaluations of their effects are lacking. Here we investigated the public health effects of a temporary traffic light system introduced in the United Kingdom (UK) in 2021, imposing red-amber-green (RAG) status based on risk assessment.

**Methods:** We analysed data on international flight passengers arriving into Scotland, COVID-19 testing surveillance, and SARS-CoV-2 whole genome sequences to quantify effects of the traffic light system on (i) international travel frequency, (ii) travel-related SARS-CoV-2 case importations, (iii) national SARS-CoV-2 case incidence, and (iv) importation of novel SARS-CoV-2 variants.

**Results:** International flight passengers arriving into Scotland had increased by 754% during the traffic light period. Amber list countries were the most frequently visited and ranked highly for SARS-CoV-2 importations and contribution to national case incidence. Rates of international travel and associated SARS-CoV-2 cases varied significantly across age, health board, and deprivation groups. Multivariable logistic regression revealed SARS-CoV-2 cases detections were less likely among travellers than non-travellers, although increasing from green-to-amber and amber-to-red lists. When examined according to travel destination, SARS-CoV-2 importation risks did not strictly follow RAG designations, and red lists did not prevent establishment of novel SARS-CoV-2 variants.

**Conclusions:** Our findings suggest that country-specific post-arrival screening undertaken in Scotland did not prohibit the public health impact of COVID-19 in Scotland. Travel rates likely contributed to patterns of high SARS-CoV-2 case importation and population impact.

## Background

On 24^th^ March 2020, Scotland with the rest of the United Kingdom (UK) entered a national lockdown to limit SARS-CoV-2 spread and subsequent COVID-19-associated hospitalisations. This early period of the pandemic was characterised by British nationals being advised against all but essential travel (1), alongside unprecedented border closures implemented increasingly by countries globally (2). However, prior to implementation of these stringent travel restrictions in the UK, hundreds of introductions of SARS-CoV-2 into Scotland had already occurred through international travel and community transmission was underway (3).

By July 2020, circulating SARS-CoV-2 was dramatically reduced (4) and Scotland had moved to Phase 3 of a route map out of lockdown (5). Whilst interventions focusing on reducing social contacts remained in place, travel restrictions were relaxed with the lifting of quarantine measures from certain overseas destinations (6). A country-specific risk assessment approach was subsequently undertaken with the introduction of ‘travel corridors’, relaxing self-isolation measures for individuals travelling to countries not considered to be high risk for high levels of SARS-CoV-2 exposure (7).

Progress out of lockdown was short lived however, with increasing COVID-19 incidence linked to travel in the summer of 2020 (4,8) and the subsequent identification of the first novel SARS-CoV-2 Variant of Concern (VOC), Alpha (B.1.1.7 Pango lineage (9)), in Scotland during November 2020, first identified in the UK and classified VOC based on risk assessment (10). Scotland subsequently entered a period with travel restrictions enforced at a local authority level based on regional infection rates, regulating both domestic and international travel, and the country entered its second national lockdown on 5^th^ January 2021. By 4^th^ January 2021, Alpha was the dominant variant circulating in the UK and managing the tradeoff between COVID-19 hospitalisations and recovery of the economy including the travel industry was of high concern. Other variants of concern were also identified, such as Beta (B.1.351) in South Africa, resulting in a long-standing travel ban to this country.

Subsequently, in line with the lifting of lockdown restrictions and safe resumption of international travel, May 2021 marked the introduction of a ‘traffic light system’ across the UK which applied relaxed quarantine and testing requirements for travellers on a country-specific basis through Red-Amber-Green (RAG) list designations (11). During this period, the epidemiological course of the epidemic shifted in the UK, with the importation of a fourth novel VOC, Delta (B.1.617.2; first identified in India)(12), although widespread rollout of the national COVID-19 vaccination programme was also in place at this time. Delta replaced Alpha as the dominant variant in Scotland by the end of May 2021 (13). The traffic light system was subsequently lifted across the UK on 4^th^ October 2021, with Scotland adopting a simplified system of ‘red list’ and ‘not red’ (14). However, a fifth novel VOC, Omicron (the BA.1 sub-lineage; first identified in South Africa), was identified in Scotland at the end of November 2021 (15,16). Several countries deemed high risk were given red list status in an attempt to slow the introduction and spread of Omicron in the UK (17), although this highly transmissible variant was soon established in the community. Travel regulations were eventually lifted incrementally and fully removed across the UK on 18^th^ March 2022 (18).

International travel continues to present the main risk pathway for importations of novel SARS-CoV-2 variants of public health significance. In the UK, a temporary increase in surveillance was recently reintroduced for individuals travelling from specific locations, owing to the evolving epidemiological situation of other nations experiencing increased COVID-19 activity and the emergence of further variants of potential public health significance (19). There is, however, a need for improved understanding of how risks of novel SARS-CoV-2 introductions and subsequent spread vary by the geography and demography of populations, to inform proportionate travel-related interventions and early warning surveillance systems. There have been few observational studies providing evidence for the public health effects of COVID-19 travel restrictions, with the majority focused on border closures put in place to curb global spread early in the pandemic (2,20,21).

Here we use Scottish community surveillance and SARS-CoV-2 whole genome sequence data to evaluate the public health effects of the COVID-19 traffic light travel policy in Scotland. Our objectives were to quantify the effect of the traffic light policy on (i) international travel frequency, (ii) temporal, demographic, and geographic patterns of SARS-CoV-2 importation risk, and the odds of SARS-CoV-2 case detections, (iii) national SARS-CoV-2 case incidence, and (iv) the risk of importing specific SARS-CoV-2 VOC across RAG groups. This evaluation has importance for providing an evidence-based approach to future public health guidance and decision making around international travel policy.

## Methods

### Datasets

#### Civil Aviation Authority data

Data on the monthly number of passenger arrivals into Scottish airports, by country and airport of origin, were provided by the Civil Aviation Authority (CAA). There were 5,508 records capturing 9,383,947 passengers in total from the period April 2019 to September 2021, from flights arriving into 11 Scottish airports from 290 airports in 70 countries. Data were grouped providing total monthly numbers of flights into Scottish airports by country of origin.

#### COVID-19 Passenger Locator Forms

From June 2020, measures were introduced requiring all UK arrivals to complete a Passenger Locator Form (PLF) to support compliance with COVID-19 travel measures (23). The dataset contained weekly data for the period 28^th^ June 2020 to 19^th^ March 2022 on the number of PLFs submitted to Border Control and the originating countries from which passengers had travelled from into Scotland. Over the 90-week period, passengers arrived into Scotland from 247 countries.

To enable like-for-like country-level comparison with PCR data, data on the Canary Islands and Madeira were merged with that of Spain and Portugal respectively in both the CAA and the PLFs.

#### COVID-19 microbiological surveillance data: Community cases

Data on SARS-CoV2 infections in Scotland were accessed from the NHS Scotland Corporate Data Warehouse (CDW), a database designed to support the analysis and reporting of COVID-19 surveillance in Scotland. The national case definition for a SARS-CoV-2 infection was applied for the study duration, whereby respiratory specimens testing positive by reverse transcription real-time Polymerase Chain Reaction (PCR) or Lateral Flow Device (LFD) tests 90 days or more after an individual’s last positive specimen were captured as a separate episode of infection (reinfection). The dataset captured demographics (age, sex, postcode location) and test information (date of sample and test result). Reasons for testing included diagnostic confirmation for those with symptoms, asymptomatic testing of close contacts to support self-isolation, and workplace testing as part of the NHS Test & Protect system.

There were 1,356,983 case records for the study period of 15^th^ February 2021 to 3^rd^ May 2022. This period spanned the introduction of post-arrival tests for travellers returning from international destinations to a period post-lifting of travel restrictions.

#### COVID-19 microbiological surveillance data: Traveller tests

The CDW database captured post-arrival travel-related tests from 15^th^ February 2021. During the traffic light period, tests were taken on or before day 2 (red, amber and green list countries) and on or after day 8 (red and amber list countries). Individuals were required to self-report the main country they travelled to when booking a COVID-19 test if they had travelled internationally within the preceding 14 days. All individuals were required to take pre-departure tests until this requirement was incrementally removed from 7^th^ January 2022.

There were 425,764 SARS-CoV-2 PCR test records from individuals reporting recent international travel over the study period. Deduplication of SARS-CoV-2 tests was conducted by grouping multiple tests based on a unique identifier (Community Health Index) for individuals with specimen dates falling within a 14-day period and relating to the same travel destination, thereby defining travel events by country. Following this process, 345,045 travel records remained representing 290,044 individuals. Over the study period, 41,934 (14.5%) of individuals had more than one travel event, with a minority of individuals (n=1,178, 0.4%) reporting travel to more than one country within a 14-day period. Tests with specimen dates out with a 90-day period were deemed a separate episode of infection (following the national case definition (24)). Positive test records were preferentially retained over negative records for each travel event. Monthly PCR test frequencies for travellers were strongly correlated with numbers of passengers into Scotland based on PLFs (Figure S1, Supplementary Material).

#### COVID-19 microbiological surveillance data: Case-control analysis

Logistic regression modelling was employed to assess the relative odds of SARS-CoV-2 case detection between international travellers and non-travellers. For this analysis, PCR tests were extracted for the traffic light period (17^th^ May 2021 to 30^th^ September 2021). There were 184,126 deduplicated travel records for this time period, and among 183,839 with the information, 1.9% had self-reported symptoms.

Non-travel records (n=4,011,152) were deduplicated as for the travel records. Those with specimen dates out with 90 days of the first identified records were presumed a separate episode of infection, retaining a single record per episode. Positive test results were retained over negative test results. Test records without a unique identifier were assumed to be from different individuals. Following deduplication, there were 1,891,873 non-travel records during the traffic light period and among 1,499,890 tests with the information, 43.3% had self-reported symptoms.

#### SARS-CoV-2 whole genome sequencing data

Whole Genome Sequencing (WGS) of SARS-CoV-2 samples was introduced in Scotland in March 2020. Following emergence of the Alpha variant, a push to sequence all positive SARS-CoV-2 samples was stepped up to allow for surveillance of variants, though with prioritisation toward travel-related samples to identify importation of VOC. WGS results were uploaded by sequencing labs and quality assessed via the COVID-19 Genomics UK (COG-UK) pipeline hosted on the Cloud Infrastructure for Microbial Bioinformatics (CLIMB) as part of COVID-19 Genomics UK (COG-UK)(25). Definitions based on mutational patterns were then applied to resultant genomic information to designate a variant (9) and these results were linked to the original PCR test via a unique specimen identifier.

During the study period there were 317,570 samples from COVID-19 cases resident in Scotland that underwent SARS-CoV-2 whole genome sequencing. Samples which had a low-quality genome and samples that could not be linked to a positive SARS-CoV-2 PCR test, and therefore could not be confirmed as a Scottish sample, were excluded (n=78).

#### Traffic light system country designations

The Red-Amber-Green (RAG) status of each travel destination was extracted from Scottish Government online travel updates (18). Test records, including genome sequences, were linked to RAG status based on the recorded travel destination and the specimen date of the test. Analyses aggregating data on a weekly basis applied the country’s RAG status pertaining to the start of each week, all other analyses applied the daily designations.

### Statistical analysis

#### Assessment of travel patterns by time, location and demography

Descriptive statistical analyses were performed to characterise key trends of international travel for the Scottish population before and during the pandemic, based on CAA and PLF data. Changes in the count of passengers entering Scotland, at or between key time points, was used as an objective indicator of the impact of travel-related measures on travel.

To assess the impact of the traffic light system on the risk of SARS-CoV-2 importations, trends in the weekly numbers and proportions of PCR-positive travellers were examined for the top 30 most frequently visited travel destinations, spanning a period before (February-May 2021), during (May 2021-October 2021), and after (October 2021-May 2022) the traffic light system. Although post-arrival tests were a mandatory travel policy, some exceptions were in place and compliance is not quantified (23). PLF data were therefore used to assess how representative the PCR test data was of travel frequency by destination during the traffic light period.

Groups at risk of importing SARS-CoV-2 through international travel were also assessed by comparing travel frequencies across demographic (age, sex, relative deprivation of residential location, as measured by the Scottish Index of Multiple Deprivation; SIMD) and geographical (territorial NHS Board) factors. An ANOVA test was used to assess differences in mean travel frequencies across demographic and geographic groups, with a p-value <0.05 applied to indicate statistical significance.

The potential impact of travel on the epidemiological situation in Scotland was assessed by quantifying the numbers of travel-related SARS-CoV-2 cases as a proportion of all Scottish SARS-CoV-2 cases. Ranking of travel destinations according to travel frequency, importation risk, and impact gave an overall assessment of risk, shown for a period of low (June 2021) and high (September 2021) travel frequency.

#### Logistic regression modelling

SARS-CoV-2 tests were extracted and deduplicated across travellers (with a recent travel event) and non-travellers (records with no recent travel event) for calendar months spanning the period of the traffic light system (17^th^ May 2021 and 30^th^ September 2021 inclusive).

The combined dataset was linked to the RAG status according to the travel destination of each record. A ‘travel status’ variable was created whereby those with an international travel event were grouped by level of RAG status and with the remaining forming a ‘non-traveller’ group.

Statistical analyses were performed to assess the association between travel status and the odds of detecting a SARS-CoV-2 case in a test-negative case-control design, adjusting for demographic, geographic and temporal factors. Unadjusted odds ratios were initially quantified in univariate binary logistic regression models examining associations between SARS-CoV-2 infection and travel status, age group (0-19y, 20-39y, 40-59y, 60-79y, 80y+), sex (male versus female), month (May, June, July, August, September) and NHS Board (fourteen territorial health boards).

Multivariable mixed-effects logistic regression modelling was then used to quantify the relative odds of SARS-CoV-2 case detection adjusting for age group, sex, and calendar month as fixed effects and NHS Board location as a random effect. Statistical interactions were assessed for all factors that were significant in the final multivariable model, with a p-value less than 0.05 indicating statistical significance. A small number of individuals (n=8990, 0.4%) had duplicate records (positive and/or negative tests). Restricting analyses to the first observed records per individual did not alter the magnitude, direction or significance of the estimated odds ratios.

All analyses were performed in R v3.6.1 (26). The glm function was applied for single effect statistical models and the lme4 (1.1-27.1 package) for mixed-effect models.

## Results

### Impact of COVID-19 policies on patterns of international travel in Scotland

Prior to the COVID-19 pandemic, numbers of international flight passengers arriving into Scotland per month were estimated to range from around 445,000 passengers (February 2019) to 952,000 (July 2019) during the peak summer period (CAA data, see Figure 1). During the COVID-19 pandemic, monthly numbers of international flight passengers decreased substantially owing to travel restrictions (Figure 1). A reduction in travel of 97.7% (to around 17 thousand passengers per month, compared to the same period in 2019) was observed during the first national lockdown (April to May, 2020), by 86.7% (to around 95 thousand passengers per month, compared to same period in 2019) during a period of relaxed travel restrictions (data from July 2020 to January, 2021), and by 87.4% (to around 111 thousand passengers per month, compared to the same period in 2019) during the period of the traffic light system (May to September 2021).

**Figure 1.**
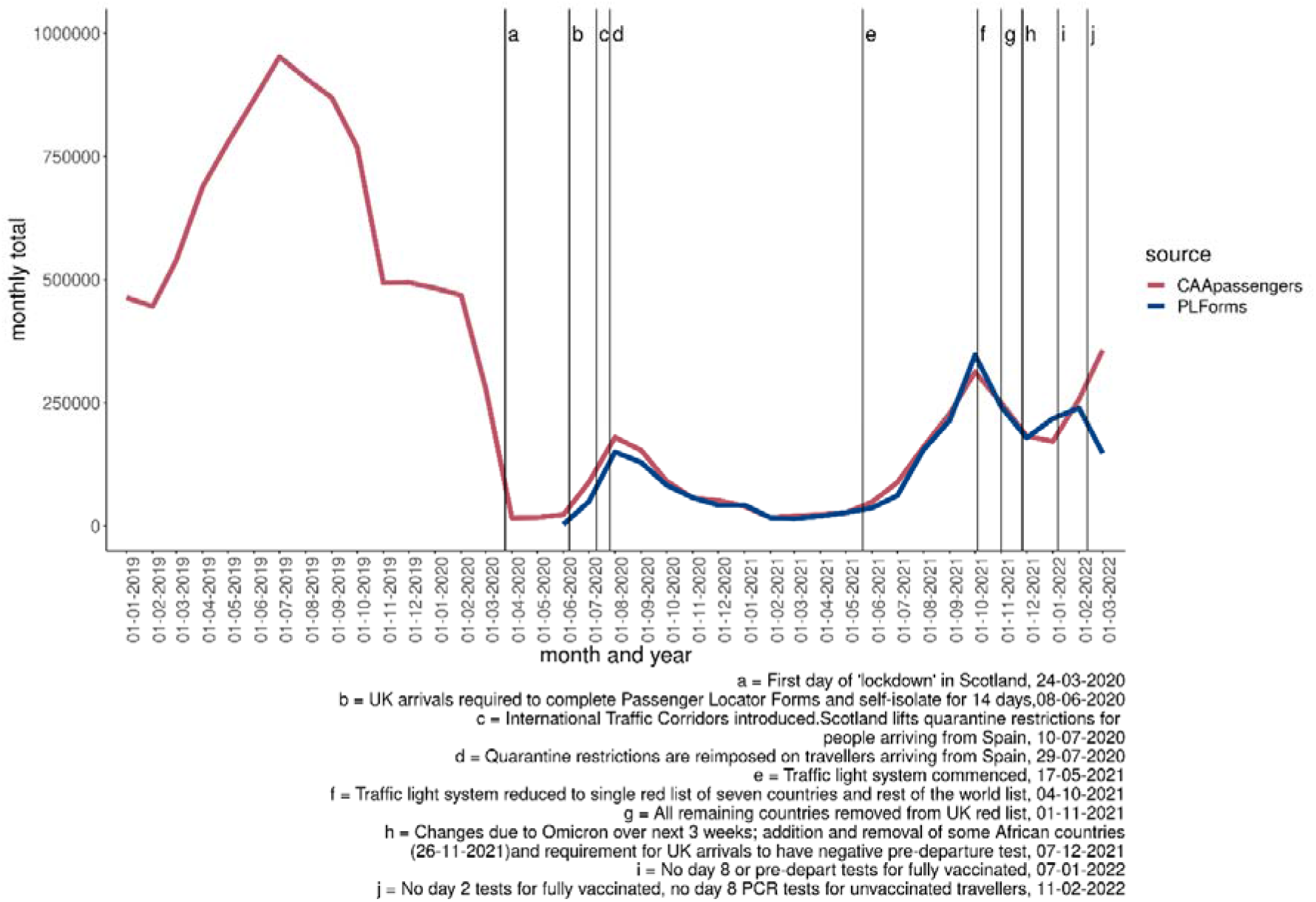
Monthly numbers of passengers into Scotland, before and during the pandemic January. Data from Civil Aviation Authority data and Passenger Locator forms spanning January 2019 to March 2022.

The magnitude of reduced international travel across the phases of COVID-19 travel restrictions was largely consistent across destinations and data sources (Figure S2, Supplementary Material). Despite large reductions in travel, the most frequently visited destinations remained largely consistent however, with Spain the most common destination both prior to and during the COVID-19 pandemic (Figure 2). The traffic light system (17^th^ May 2021 to 4^th^ October 2021) followed a country-specific risk-based approach (see Figure S3, Supplementary Material), with quarantine reserved for high-risk countries (red list), and others requiring self-isolation together with day 2 and 8 post arrival tests (amber list), or day-8 post-arrival tests only (green list). The frequency of international arrivals into Scottish airports subsequently increased by 754% from May 2021 to September 2021, compared with a 12% increase over the same period in 2019 (Figure 2).

**Figure 2.**
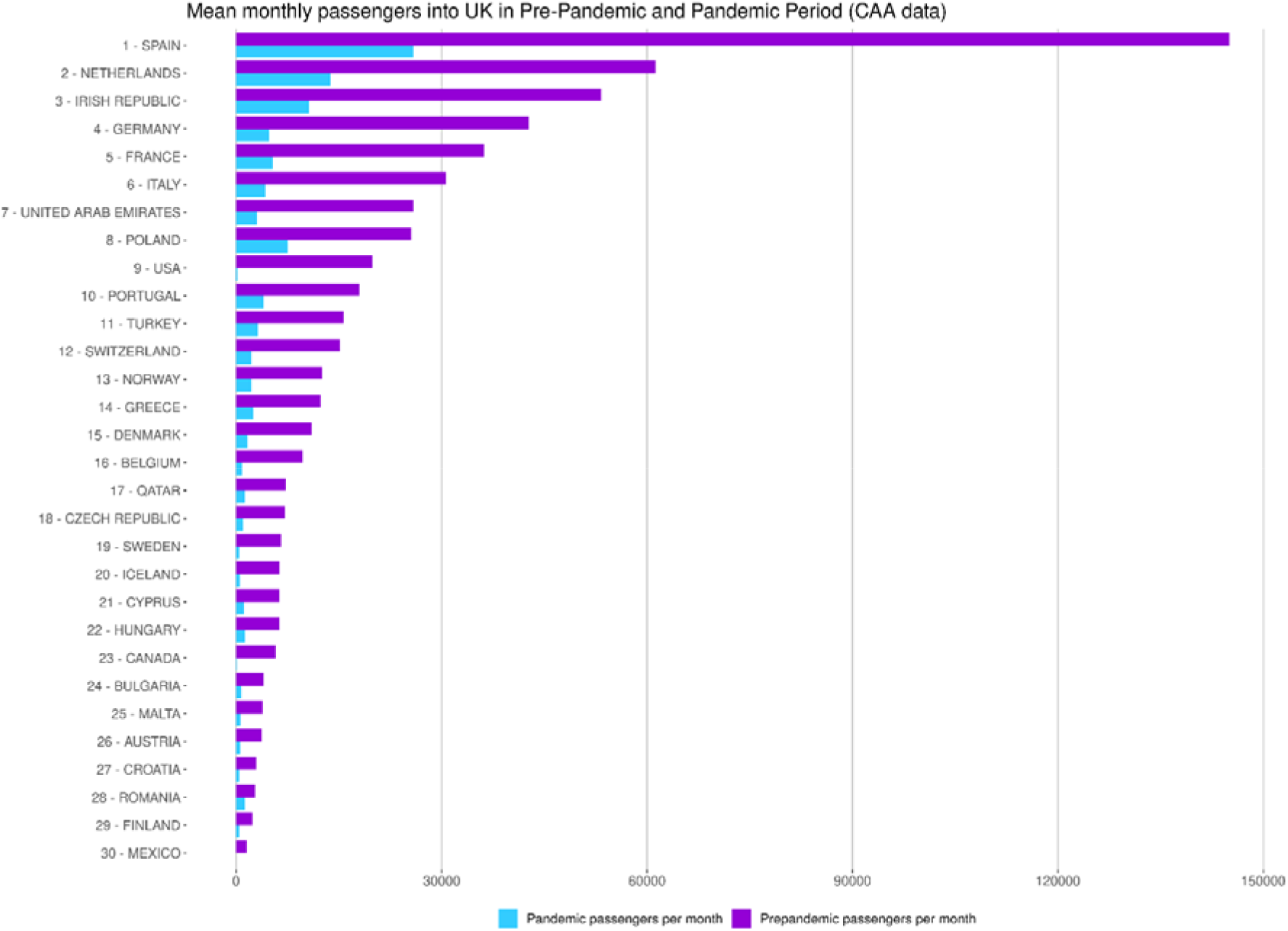
Mean monthly number of passengers with Scottish residence travelling into the UK. Based on Civil Aviation Authority data comparing periods before (January 2019 to February 2020) and during (March 2020 – March 2022) the COVID-19 pandemic.

The ranking of travel frequency across RAG groups remained consistent over time when retrospectively applying designations to the pre-pandemic period, with amber list countries the most frequently visited (83.8% and 70.0% in the pandemic period based on PLF and CAA data sources respectively, and 74.8% pre-pandemic (CAA data)). A small discrepancy was seen between red and green list countries although they were largely consistent throughout the traffic light period (Figure S3, Supplementary Material). The largest decrease in travel frequency, comparing May-to-September inclusive pre-and-post pandemic, was seen for red list (87.7% decrease) followed by amber (78.8% decrease) and green list (63.2% decrease) countries (Figure S2, Supplementary Material).

Despite the lifting of the traffic light system and incremental removal of travel restrictions in winter 2021/22, the frequency of international travel declined during this period consistent with the typical seasonality of travel (Figure 1).

### Travel and case patterns by demographic and geographic groups

A significant difference in average weekly rates of travel was found by age group, deprivation, and NHS Board, but not for sex, with a similar pattern observed for SARS-CoV-2 detection rates among travellers (Table 1). Both travel and SARS-CoV-2 case detection rates were notably higher in working aged adults (age groups 20-39y and 40-59y), for residents of NHS Boards with larger populations, and increased with decreasing deprivation (over half of all travel events were from individuals residing in the lowest deprivation groups).

**Table 1.**
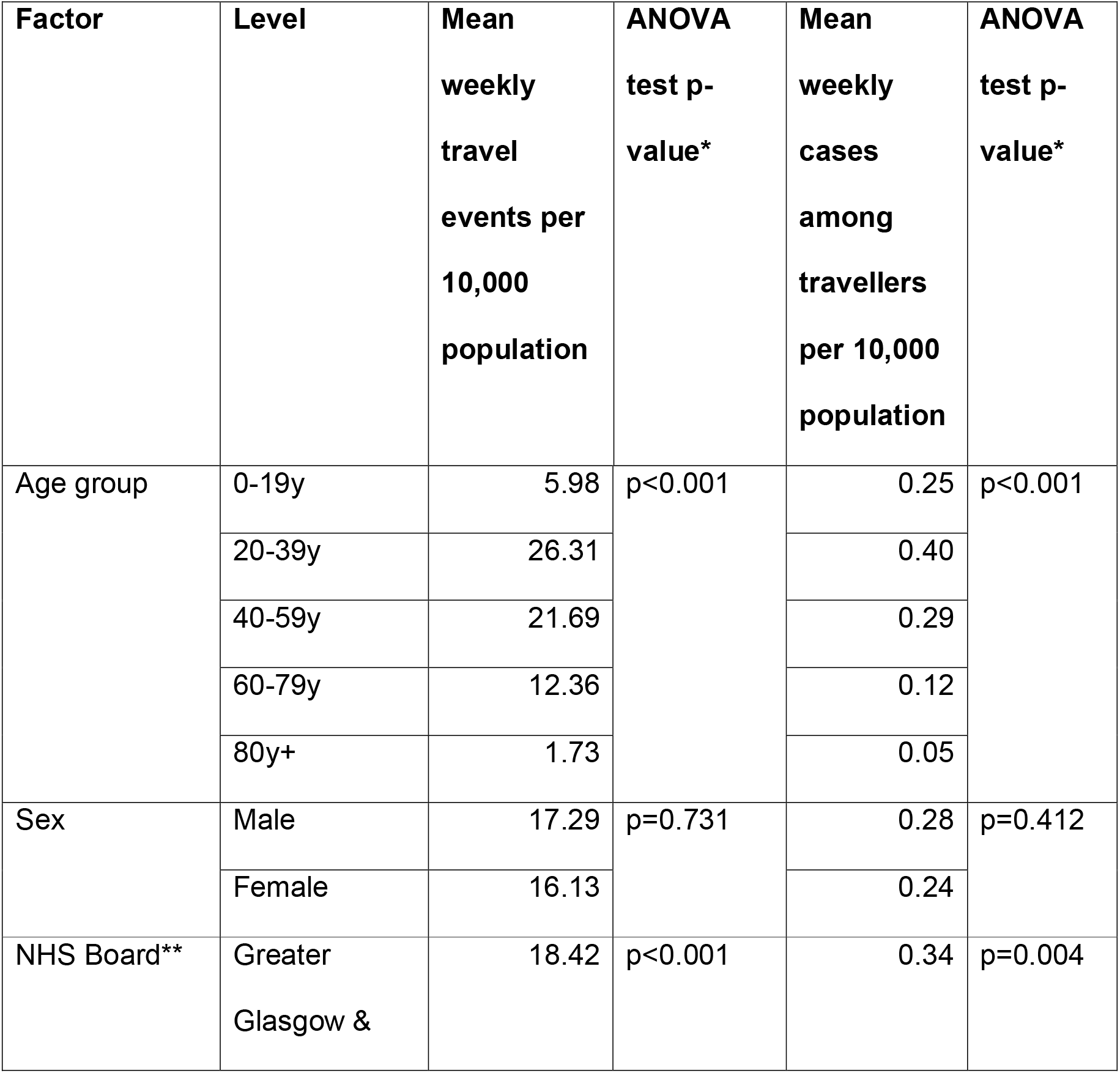
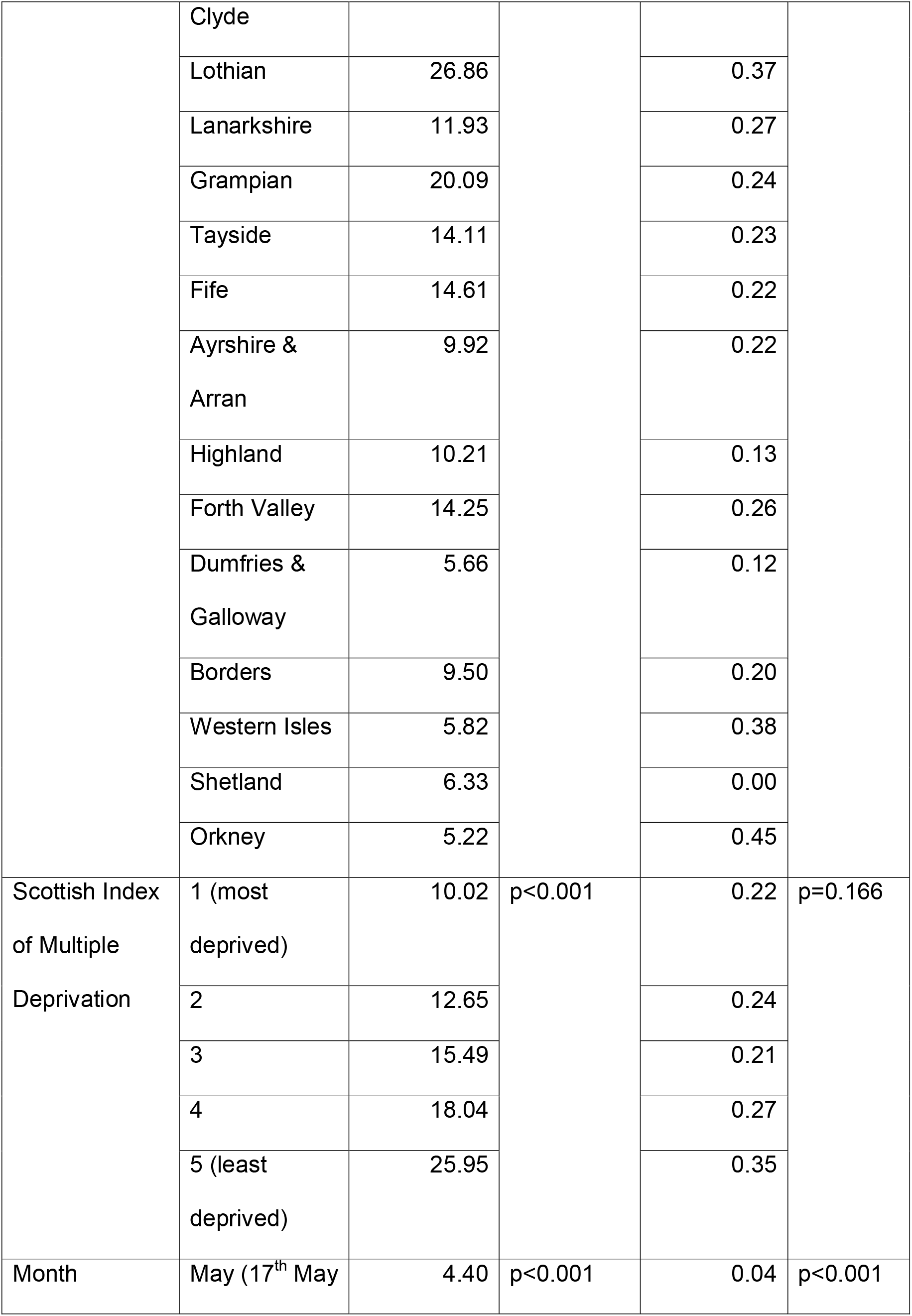
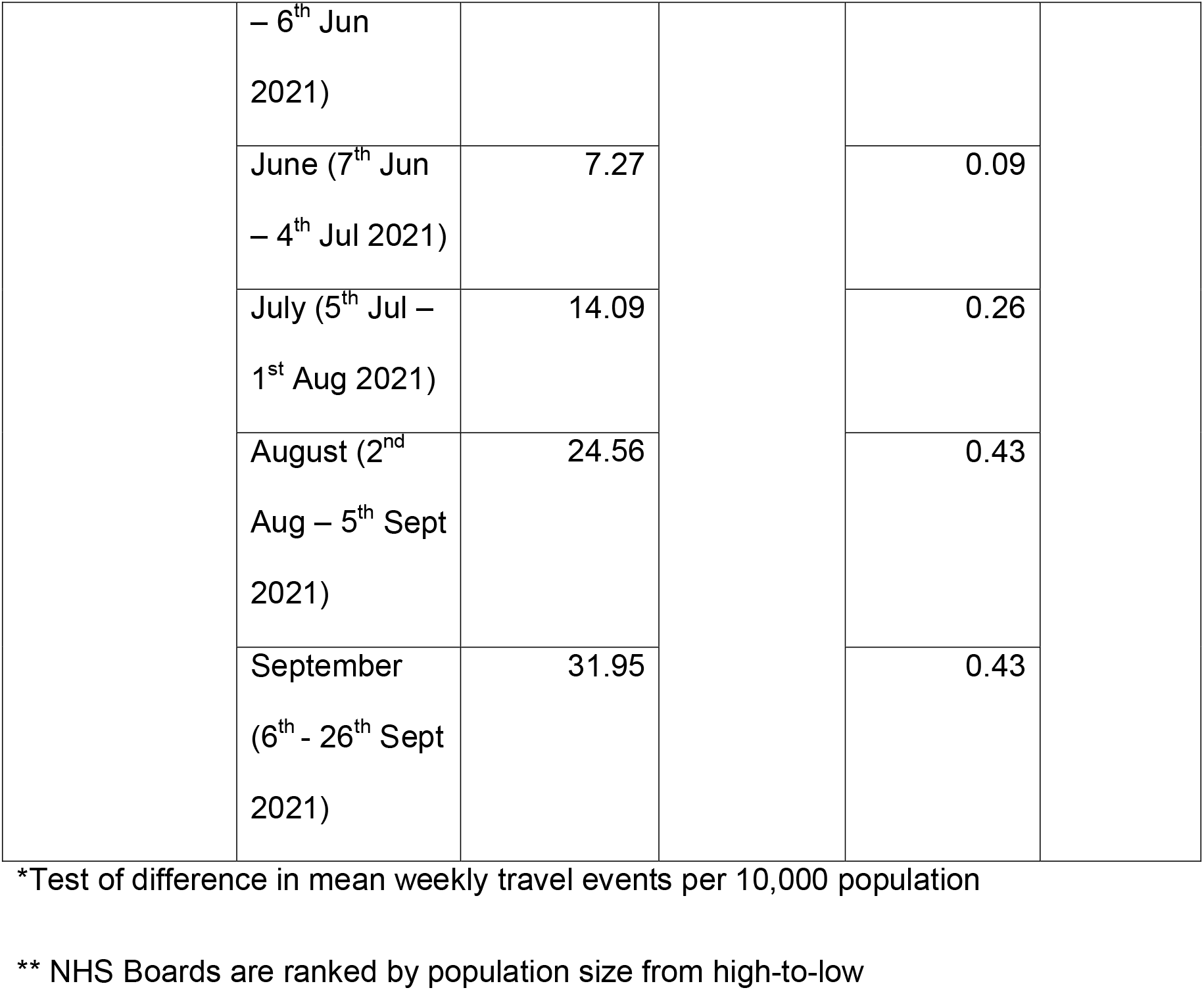
Frequency of international travel events across demographic and geographical groups, during the period of the traffic light system (17th May 2021 to 26th September 2021) in Scotland.

However, some variation was seen, with younger (20-29y) working aged females and older (30-39y and 50-59y) working aged males having the greatest travel frequencies (Figure S4a), and a relatively high proportion of travellers resided in high deprivation (low SIMD score) locations for specific destinations (Figure S4b). Despite the temporal variation, differences across NHS Boards showed general consistency with more populous regions tending to have higher travel rates (Figure S4c).

### Risks and population impact of SARS-CoV-2 importations through international travel during the period of the traffic light system

Figure 3 summarises weekly numbers of tests and travel-related SARS-CoV-2 cases for each of the top 30 most frequently visited travel destinations. Overall, there was a 324% increase in SARS-CoV-2 cases comparing the weeks with the highest travel frequency in the pre-traffic light (w/c 5^th^ April 2021) and traffic light (w/c 13^th^ September 2021) periods. For some countries, periods of increasing case numbers appeared to coincide with increasing travel frequency. However, in other instances, increasing case numbers appeared decoupled from travel frequency and instead appeared to be explained by SARS-CoV-2 importation risk (the proportion of traveller PCR tests with a positive result).

**Figure 3.**
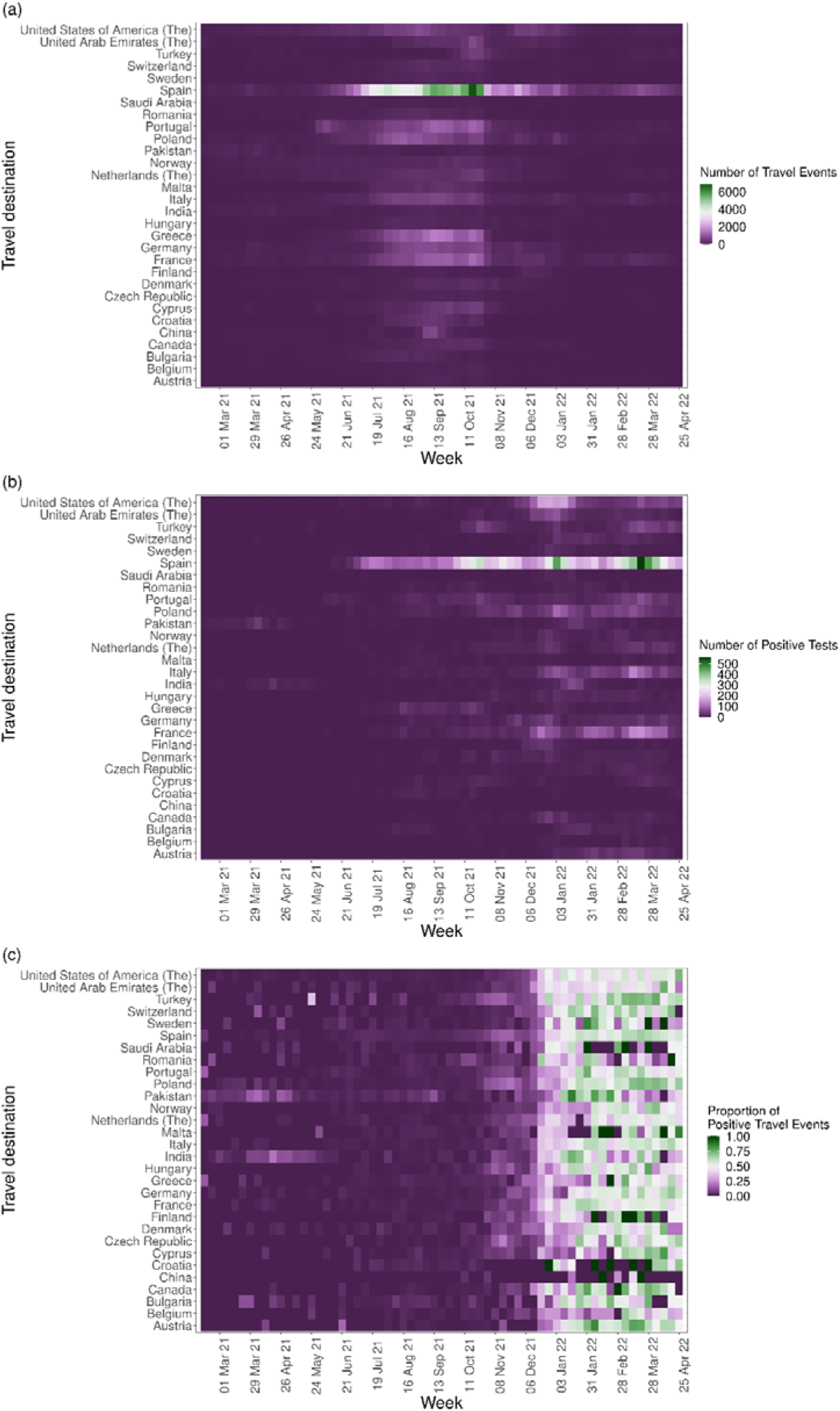
Patterns of international travel and associated SARS-CoV-2 infections in Scotland. Weekly numbers of (a) international travel events among PCR-tested individuals, (b) proportions testing SARS-CoV-2 positive, and (c) numbers testing SARS-CoV-2 positive, spanning w/c 15^th^ February 2021 to w/c 25^th^ April 2022.

Figure 4 summarises the rankings of the top 30 most frequently visited countries according to travel frequency, SARS-CoV-2 importation risk, and population impact (as measured by the proportion of all Scottish SARS-CoV-2-positive PCR tests with an associated travel event). Countries ranking high across each metric represented the greatest risk when combining exposure and consequence. Notably, four red list countries appeared in the top seven, and two in the top three, ranking of SARS-CoV-2 importation risks during the lowest (June) and highest (September) travel frequency periods respectively. However, one green list country had the highest combined ranking across all metrics during June, and by September there was no discernible pattern of risk distinguishing green and amber list destinations.

**Figure 4.**
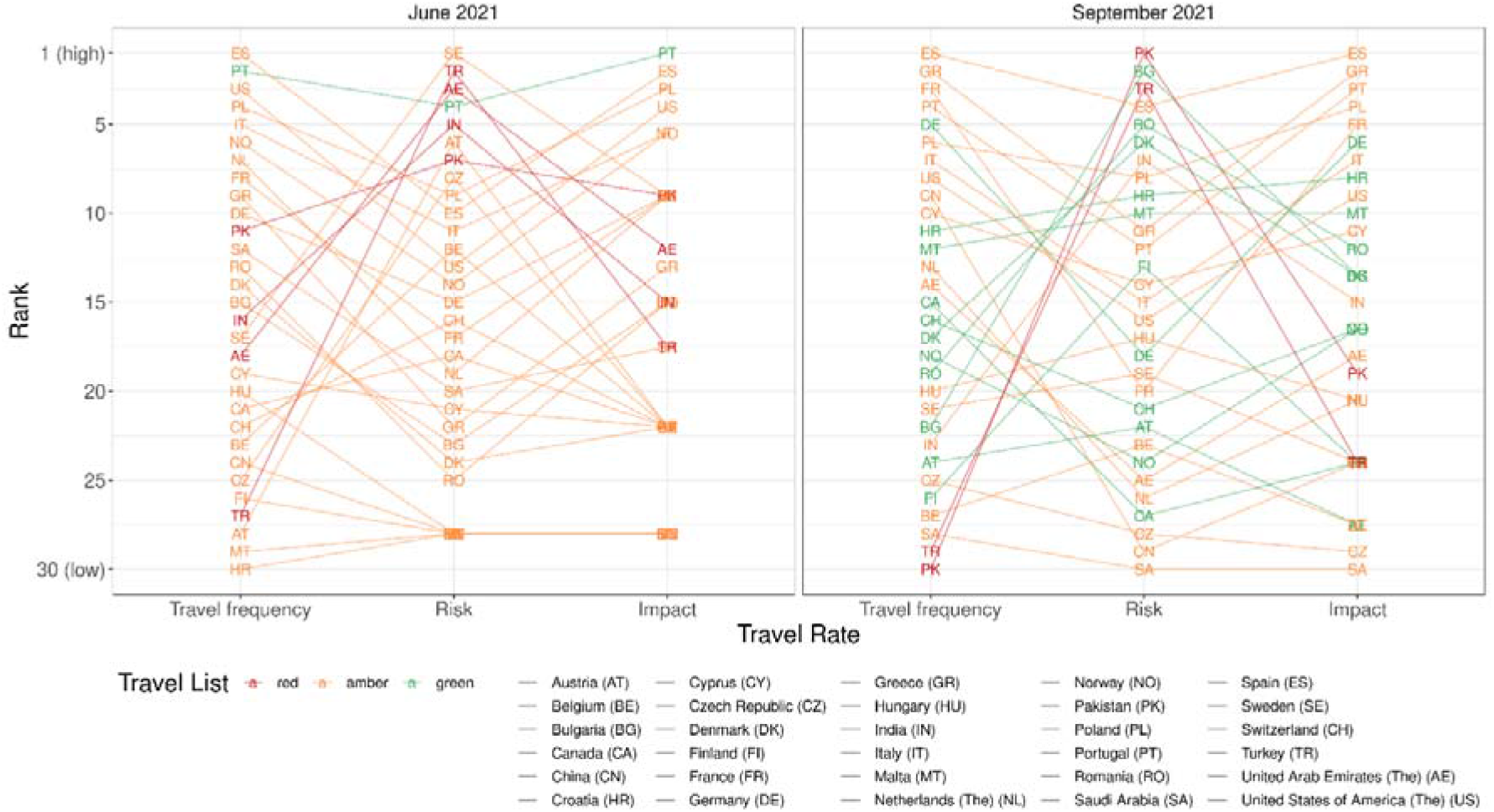
Top 30 most frequently visited international travel destinations for Scotland by risk and impact. Travel frequency = the number of travel events to the given destination; Risk = infection importation risk (as measured by the proportion of PCR-confirmed SARS-CoV-2 cases among travel-related tests); Impact = epidemiological impact (as measured by the proportion of Scottish cases of SARS-CoV-2 associated with international travel).

**Figure 5.**
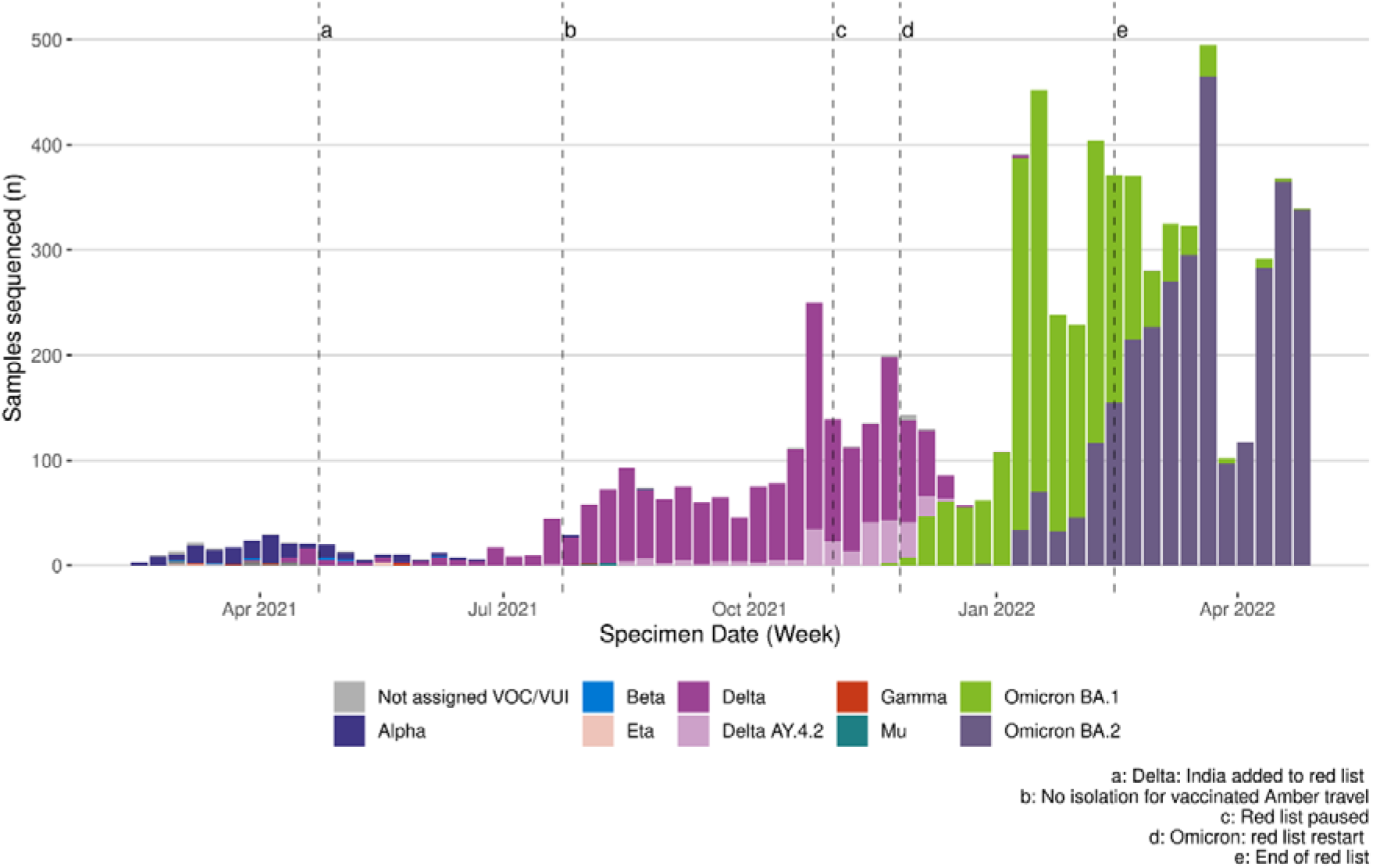
Weekly count of PCR-confirmed cases undergoing whole genome sequencing by SARS-CoV-2 variant type.

Overall, the rate of SARS-CoV-2 cases detections was estimated to be 17 per 1,000 among those with an international travel event, compared to 190 per 1,000 among those without an international travel event over the same period. Employing multivariable mixed-effects logistic regression, the odds of detecting SARS-CoV-2 cases increased from green-to-amber and amber-to-red list countries but was lower for travellers relative to the non-traveller PCR-tested community. This finding was consistent regardless of RAG group, controlling for age group, sex, month and geographical location (see Table 2 for details). Some variation in the effect of travel status was found according to age group, sex, and month, however the relative odds of case detection was consistently greater among those with no recorded recent international travel event.

**Table 2.**
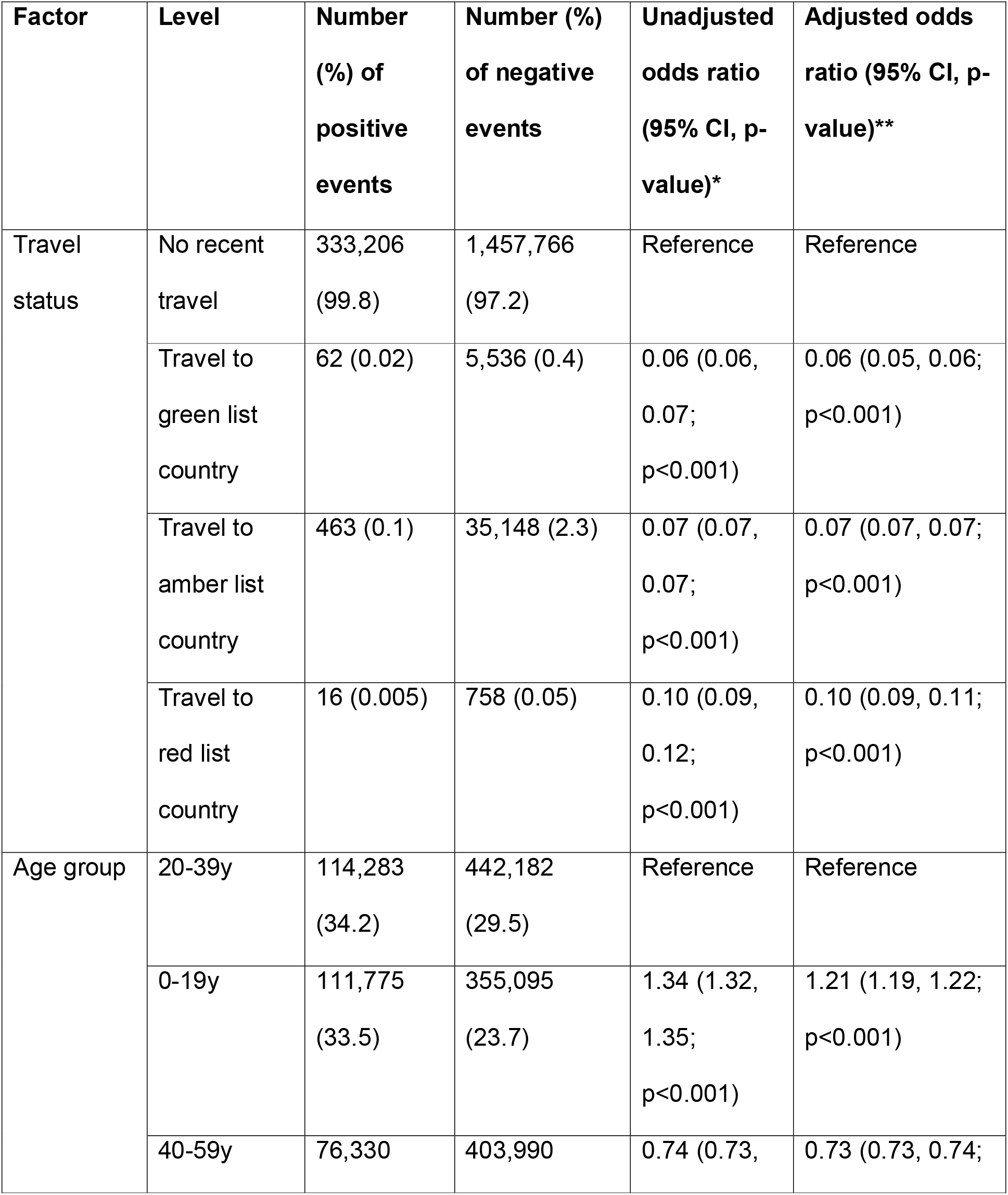
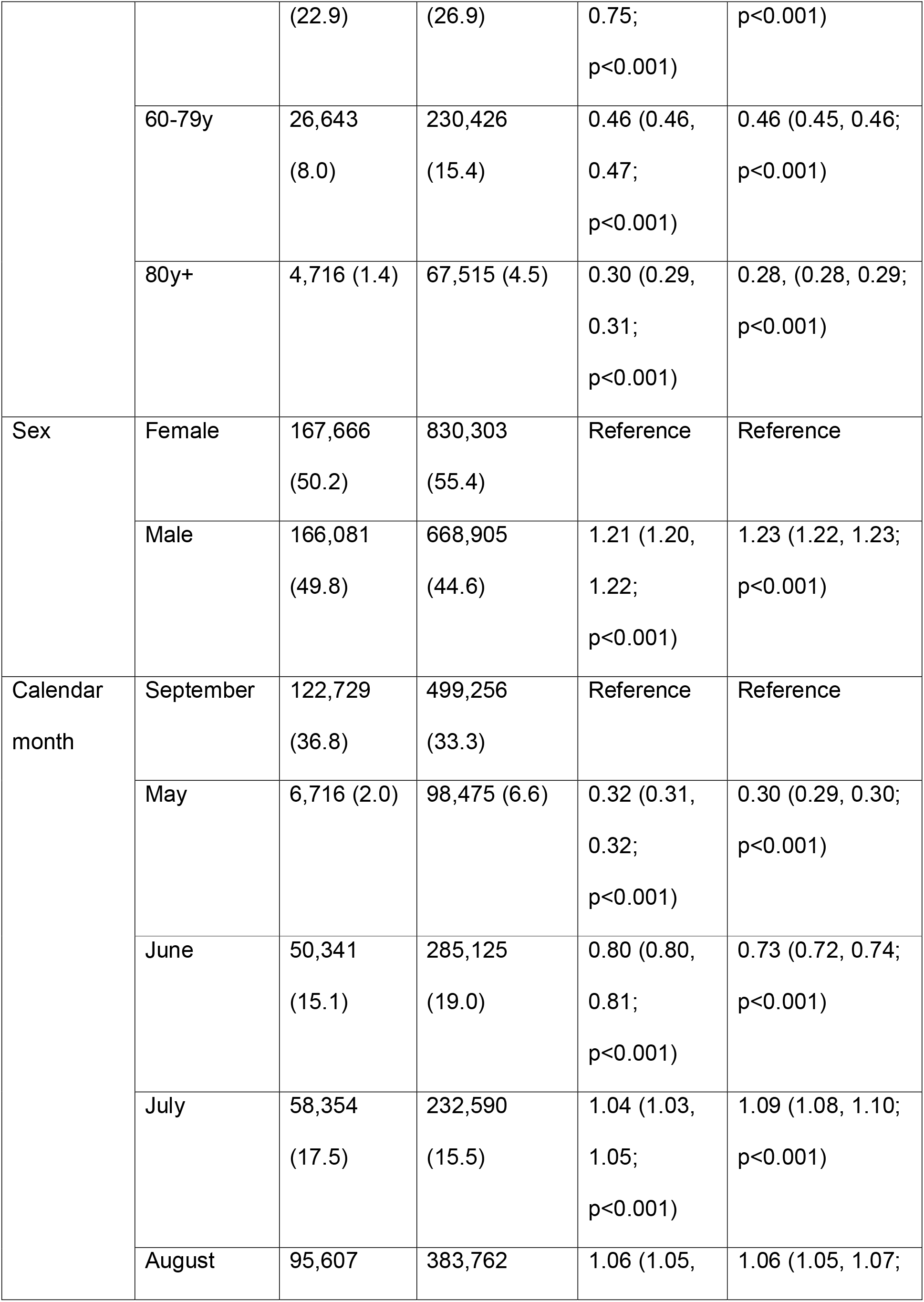
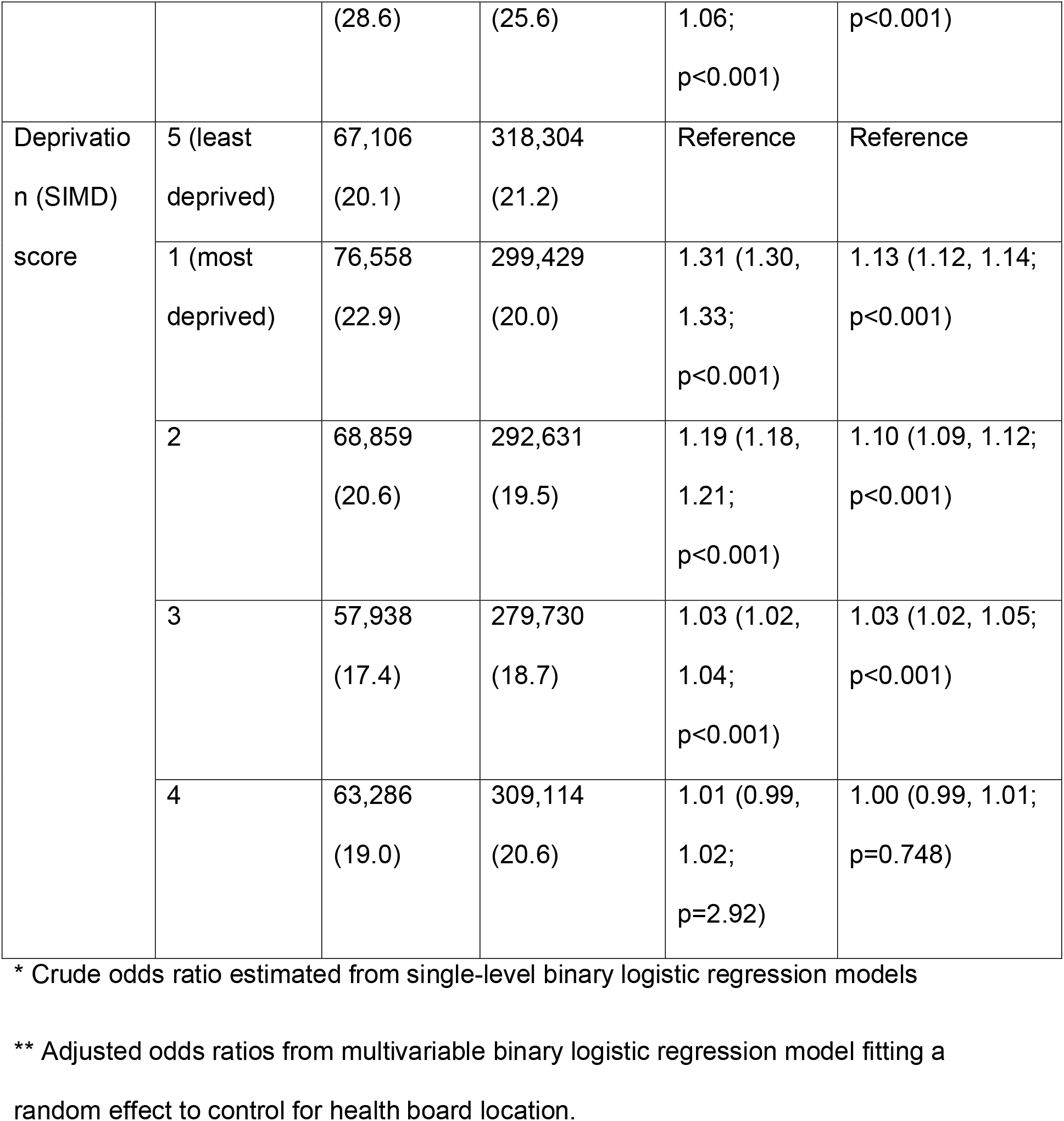
Investigating risk factors for SARS-CoV-2 infection during the period of the traffic light system (17th May 2021 to 30th September 2021) in Scotland.

Several demographic factors were identified as independently associated with the odds of detecting SARS-CoV-2, regardless of travel status, with cases more likely detected among 0-19y olds relative to 20-39y olds, among males relative to females, and among individuals’ resident in more deprived areas. These predictors of SARS-CoV-2 cases differed from that observed among travellers (as highlighted also by differences in the modelled predicted probabilities when comparing traveller and non-traveller groups; results not shown) and the patterns of travel frequency.

### Risks and population impact of SARS-CoV-2 novel variant importations through international travel during the period of the traffic light system

Figure 5 shows the weekly numbers of sequenced SARS-CoV-2 cases by variant group against a timeline of travel policy changes. For most travel destinations and periods of the epidemic, the most likely imported VOC reflected the dominant variant circulating in the UK representing 50% or more of sequenced cases (Figure S5, Supplementary Material). However, some variation in the risk of VOC importation was observed across travel destinations, particularly during the period of Alpha variant dominance albeit based on small numbers of sequenced cases.

The Delta variant was identified in Scotland on 4^th^ April 2021 before the relaxation of travel restrictions for some destinations through the traffic light system. Figure 6 compares weekly numbers of variants identified among sequenced SARS-CoV-2 cases returning from non-red and red-list travel destinations, alongside non-traveller community cases during the period in which Delta emerged. Although several countries were added to the red list from 9^th^ April 2021, Delta was primarily detected among travellers returning from non-red list countries (except for a short period from late April to end of May). Community transmission was evident from late-April, following which Delta was relatively more frequently identified among non-travellers. Delta replaced Alpha to become the dominant variant in Scotland from 19^th^ May 2021 (Figure S6, Supplementary Material).

**Figure 6.**
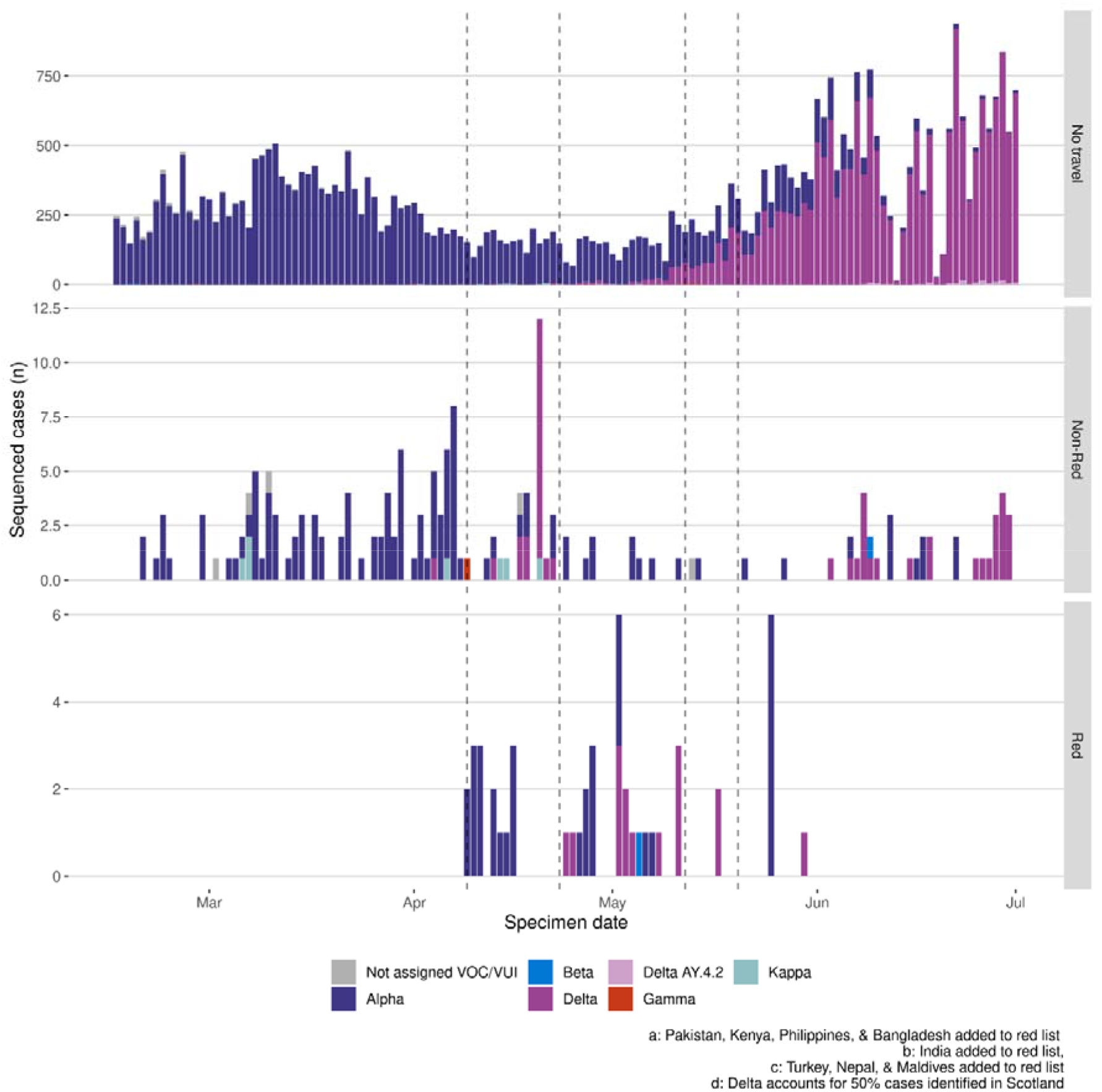
**Weekly count of whole genome sequenced SARS-CoV-2 cases during period of Delta variant emergence**. Numbers are shown among non-traveller community cases (top panel), travellers returning from non-red destinations (middle panel), and travellers returning from red-list travel destinations (bottom panel).

Shortly after the traffic light system was removed on 4^th^ October 2021, Omicron (BA.1 sublineage), emerged with the specimen of the first known case in Scotland dated 22^nd^ October 2021 (Figure S6, Supplementary Material). In contrast to Delta, Omicron was first identified in the community among non-travel-related cases (Figure 7), with only 2.6% of sequenced Omicron cases associated with recent international travel compared to 22.1% of Delta cases in the first four weeks of their respective emergence. Overall, both Delta and Omicron were more frequently detected among non-travellers and similarly to Delta, detection frequency for Omicron was higher in those returning from non-red list than red list countries.

**Figure 7:**
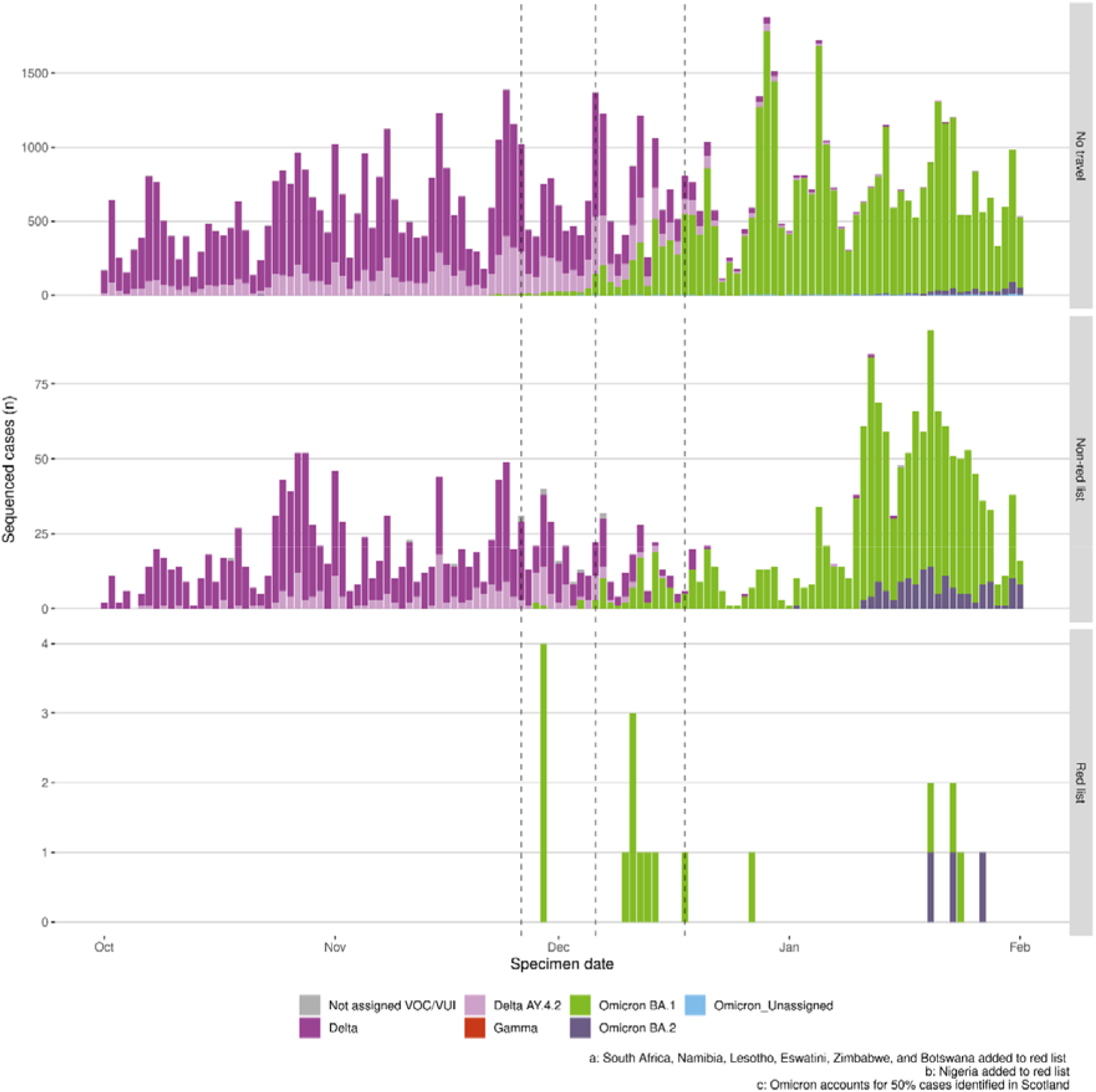
Weekly count of whole genome sequenced SARS-CoV-2 cases during period of Omicron variant emergence. Numbers are shown among non-traveller community cases (top panel), travellers returning from non-red destinations (middle panel), and travellers returning from red-list travel destinations (bottom panel).

## Discussion

With the ongoing generation of novel variants able to infect vaccinated and previously infected individuals, SARS-CoV-2 can be expected to persist globally with continued spread of novel variants of public health significance. While acquired immunity either from prior waves of infection or from vaccines are now mediating disease severity in most cases of infection, the future evolution of SARS-CoV-2 remains unpredictable. In the event of a new VOC emerging, border closures may provide a means for reducing the numbers of imported cases and control the progression of outbreaks locally. However, it is well recognised that unless implemented rapidly and informed by the true risks, the public health benefit is limited.

Once community transmission has been established, risk-based quarantine and post-travel screening may be proportionate measures to reduce the public health consequences of international travel. Risk-based screening measures have recently been reintroduced by several nations, including the UK, owing to increasing global COVID-19 spread (19). However, there is a lack of understanding of travel policy effectiveness pre-dating COVID-19, including for pandemic influenza and other emerging infectious diseases (27,28). Most studies assessing the public health effects of COVID-19 travel-related policies have used modelling approaches in the absence of empirical data, and have focused on the early phase of the pandemic when reduced international travel was of paramount interest to curb the global spread of disease (2,20,21,29,30). One study suggests the quarantine of symptomatic COVID-19 cases is insufficient to significantly delay the onset of an epidemic (31).

The traffic light system employed in the UK during May to October 2021 aimed to provide a risk-based approach to targeting quarantine and screening measures for travellers returning from specific destinations. To our knowledge, our study is the first to evaluate the potential public health effects of this measure. Firstly, our study shows, as expected, that international travel increased substantially during the period of the traffic light system. The highest travel rates were seen for working aged adults, and those resident in more populous health boards and in low deprivation areas. The ability to characterise patterns of travel may have implications for targeting safe travel messaging in the prevention of future VOC importations.

Secondly, travel was most frequent to amber list countries during the traffic light period despite the mandatory requirement for self-isolation. This finding was corroborated across PLF and CAA datasets despite known limitations, with the former involving self-submitted information and therefore reliant on compliance, and with the latter expected to have comparatively underrepresented travel from countries with few or no direct flights into Scotland but overrepresented travel from airports operating as central hubs.

Thirdly, there were significant differences in the rates of SARS-CoV-2 infections detected amongst travellers across age groups and deprivation, but not for sex, aligning with patterns of travel frequency. However, cases of SARS-CoV-2 were more likely detected among non-travellers than travellers overall during the traffic light period, and the relative odds of detecting cases differed from the pattern of cases observed among travellers. Overall these findings highlight the complexity in understanding predictors of case detections that are specific to travellers, but that patterns of travel may be a good proxy indicator of this.

The likelihood of detecting a SARS-CoV-2 infection among travellers increased from green-to-amber-to-red list countries, although with some variation when examining individual travel destinations. The highest frequency of travel was seen for an amber list country, resulting in relatively high numbers of imported SARS-CoV-2 cases when coupled with importation risk (proportion of travellers testing positive) together with a high population impact (proportion of Scottish SARS-CoV-2 cases attributed to travel). However, our study shows that despite fewer travel events, the highest SARS-CoV-2 importation risk was associated with a green list country in June 2021. Furthermore, by September 2021, a number of green list countries ranked higher than red list countries for population impact, highlighting the complexity of proportionate applications of RAG systems. We note however that our study does not assess the impact of quarantine and isolation measures in place for those returning from red and amber list countries which is expected to have reduced the population impact of international travel. Travel frequency and the context of existing community circulation are key factors affecting the public health effects of international travel, along with the epidemiological situation of the travel destination. Identifying specific countries as high risk without considering the speed of spread of SARS-CoV-2 variants to other regions internationally, coupled with large differences in international surveillance efforts, will severely limit the effect of a traffic-light system.

Finally, our study spanned two novel SARS-CoV-2 variant introductions. Stringent travel restrictions did not prevent the introduction of Delta into the UK, whilst the risk-based red list policy did not prevent establishment in the community once introductions had occurred. However, decreased case numbers were observed for some red list countries during the Delta wave despite a high SARS-CoV-2 importation risk, likely owing to decreased travel to these destinations. The rapidity by which community transmission was established in the case of Omicron, with most initial cases detected among non-travellers, suggests the direct impact of travel to red list countries during this period was likely minimal, especially given the requirement for quarantine. It was not possible however to assess the impact of the evolving epidemiological situation in red list countries and degree of global spread on the risks of Omicron variant importations from amber and green lists countries.

Our study has a number of limitations. Firstly, laboratory surveillance data may not capture all travellers; an unknown number of individuals were exempt from testing and compliance is not quantifiable. Secondly, deductions of SARS-CoV-2 infection risks across traveller and non-traveller groups must be made with caution. The greater odds of detecting SARS-CoV-2 cases in the non-traveller group may be explained by the targeted testing of symptomatics and known close contacts of confirmed cases.

Furthermore, those entering the UK were required to take a pre-departure test during the traffic light period, so the proportion of cases in this group is expected to reflect the risk of SARS-CoV-2 importation - combining the risks of a travel-associated infection and testing negative prior to departure. This should not preclude the validity of comparing SARS-CoV-2 case frequency over time, by travel destination, and across demographic and geographic groups. Thirdly, case misclassifications may have arisen, with some over-ascription of infections to the period of international travel (the acquisition of infection before or after travel cannot be ruled out, including from household transmission). Fourthly, in the absence of a suitable control population or period, our study did not assess the reduction in SARS-CoV-2 case incidence in Scotland attributable to the traffic light system.

## Conclusions

Our findings show that risk-based post-arrival screening undertaken in Scotland did not in practice prohibit the importation of SARS-CoV-2 cases, or the establishment of SARS-CoV-2 VOC in the Scottish community, arising through international travel. Overall SARS-CoV-2 case importation risks did not strictly follow RAG designations. This is likely explained by travel frequency, regardless of country risk status, coupled with rapid global spread and local community transmission limiting the value of red lists to control VOC importations and population impact. Our study provides valuable learnings that may support decision making in Scotland during any future emergency response for COVID-19 or pandemics. These lessons may be of relevance to other nations and infectious diseases although differences in the epidemiology, disease surveillance systems, and travel policy implementations should be borne in mind. While it is clear that risk-based travel restrictions may have limited value in isolation in the case of a pathogen like SARS-CoV-2, which has the potential to spread rapidly, they could potentially work more effectively if international variant surveillance systems were compatible, and with information shared in a timely and open manner, to rapidly detect and respond to new variants as they arise.

## Declarations

### Ethics approval and consent to participate

This study was undertaken as part of public health surveillance activity within the COVID-19 programme of Public Health Scotland, in line with the necessary associated regulations and guidelines. The retention and processing of information on individuals is conducted by Public Health Scotland as part of COVID-19 surveillance in Scotland in the context of emergency data processing (https://www.informationgovernance.scot.nhs.uk/covid-19-privacy-statement/), including the Civil Contingencies Act 2004, the NHS (Scotland) Act 1978 and the Public Health (Scotland) Act 2008, and under Articles 6(1)(e), 9(2)(h), 9(2)(i), 9(2)(j) of the General Data Protection Regulation. Surveillance data was shared with NHS Scotland according to the Intra NHS Scotland Data Sharing Accord (https://www.informationgovernance.scot.nhs.uk/wp-content/uploads/2020/06/2020-06-17-Intra-NHS-Scotland-Sharing-Accord-v2.0.pdf). Ethics approval and informed consent was not required for this work which was based on pre-existing infectious disease surveillance data for the Scottish population. The access and processing of individual-level data was conducted under standard Data Protection Impact Assessment information governance approval. Individual-level records were de-identified before being accessed and analysed by the project team.

## Consent for publication

Not applicable.

## Availability of data and materials

The healthcare data used in this study are available upon application to the NHS Scotland Public Benefit and Privacy Panel For Health and Social Care: <u>Public Benefit and Privacy Panel for Health and Social Care (scot.nhs.uk)</u>. International flight data may be sourced from the Civil Aviation Authority (https://www.caa.co.uk/home/); see also: https://www.caa.co.uk/data-and-analysis/uk-aviation-market/airports/uk-airport-data/.

## Authors’ contributions

SL, MTG, DLR, ASP, JH and SN conceived the study and acquired funding. IM, SH, KL, SL, JH and SN designed the analyses. JB performed data extractions and processing. IM, SH, KL, GY, SS and NC performed data curation. IM, SH, and KL performed analyses. MH, DLR, ASP, JH and SN coordinated data generation. SN coordinated and supervised analyses and the study team. All authors interpreted data. IM, SH, KL and SN wrote the first draft of the manuscript. All authors contributed to and approved the final version.

## Data Availability

The healthcare data used in this study are available upon application to the NHS Scotland Public Benefit and Privacy Panel For Health and Social Care: https://www.informationgovernance.scot.nhs.uk/pbpphsc/. International flight data may be sourced from the Civil Aviation Authority (https://www.caa.co.uk/home/); see also: https://www.caa.co.uk/data-and-analysis/uk-aviation-market/airports/uk-airport-data/.

## Acknowledgments

We thank the Bioinformatics, COVID-19 Surveillance, and Travel Health teams at Public Health Scotland who developed and managed the surveillance infrastructure and data sharing that enabled this study as part of the COVID-19 response in Scotland. We also thank the Civil Aviation Authority, in particular Kit Beynon, for providing the data analysed in our study and for their helpful comments. We also thank Jim McMenamin for a helpful critique of the manuscript. The SARS-CoV-2 genomics data was initially generated through a network of laboratories as part of the COVID-19 Genomics UK Consortium (<u>COVID-19 Genomics UK Consortium (cogconsortium.uk)</u>) and then by the NHS Sequencing Service in Scotland (<u>COVID-19 Whole Genome Sequencing (WGS) - Public health microbiology - Services - Public Health Scotland</u>).

## References

1. Coronavirus (COVID-19) update: First Minister’s speech 24 March 2020 - gov.scot [Internet]. [cited 2022 Oct 7]. Available from: https://www.gov.scot/publications/first-ministers-update-covid-19/

2. Bou-Karroum L, Khabsa J, Jabbour M, Hilal N, Haidar Z, Abi Khalil P, et al. Public health effects of travel-related policies on the COVID-19 pandemic: A mixed-methods systematic review. J Infect. 2021 Oct;83(4):413–423.

3. da Silva Filipe A, Shepherd JG, Williams T, Hughes J, Aranday-Cortes E, Asamaphan P, et al. Genomic epidemiology reveals multiple introductions of SARS-CoV-2 from mainland Europe into Scotland. Nat Microbiol. 2021 Jan;6(1):112–122.

4. Lycett SJ, Hughes J, McHugh MP, Filipe ADS, Dewar R, Lu L, et al. Epidemic waves of COVID-19 in Scotland: a genomic perspective on the impact of the introduction and relaxation of lockdown on SARS-CoV-2. medRxiv. 2021 Jan 20;

5. Coronavirus (COVID-19) Phase 3: Scotland’s route map update - 9 July 2020 - gov.scot [Internet]. [cited 2021 Oct 10]. Available from: https://www.gov.scot/publications/coronavirus-covid-19-framework-decision-making-scotlands-route-map-through-out-crisis-phase-3-update/

6. Quarantine rule ends for travellers arriving from lower risk countries and territories - gov.scot [Internet]. [cited 2022 Oct 6]. Available from: https://www.gov.scot/news/quarantine-rule-ends-for-travellers-arriving-from-lower-risk-countries-and-territories/

7. [Withdrawn] COVID-19 risk assessment methodology for inbound international travel - GOV.UK [Internet]. [cited 2022 Nov 28]. Available from: https://www.gov.uk/government/publications/covid-19-risk-assessment-methodology-for-inbound-international-travel/covid-19-risk-assessment-methodology-for-inbound-international-travel

8. Hodcroft EB, Zuber M, Nadeau S, Vaughan TG, Crawford KHD, Althaus CL, et al. Spread of a SARS-CoV-2 variant through Europe in the summer of 2020. Nature. 2021 Jul;595(7869):707–712.

9. O’Toole Á, Scher E, Underwood A, Jackson B, Hill V, McCrone JT, et al. Assignment of epidemiological lineages in an emerging pandemic using the pangolin tool. Virus Evol. 2021 Jul 30;7(2):veab064.

10. Hill V, Du Plessis L, Peacock TP, Aggarwal D, Colquhoun R, Carabelli AM, et al. The origins and molecular evolution of SARS-CoV-2 lineage B.1.1.7 in the UK. Virus Evol. 2022 Aug 26;8(2):veac080.

11. Traffic light system: safe return to international travel - GOV.UK [Internet]. [cited 2022 Nov 28]. Available from: https://www.gov.uk/government/speeches/traffic-light-system-safe-return-to-international-travel

12. McCrone JT, Hill V, Bajaj S, Pena RE, Lambert BC, Inward R, et al. Context-specific emergence and growth of the SARS-CoV-2 Delta variant. Nature. 2022 Oct;610(7930):154–160.

13. Tracking the Delta variant in Scotland - Our blog - Public Health Scotland [Internet]. [cited 2022 Oct 6]. Available from: https://www.publichealthscotland.scot/our-blog/2021/november/tracking-the-delta-variant-in-scotland/

14. Summary of updates to international travel, October 2021 - GOV.UK [Internet]. [cited 2022 Oct 11]. Available from: https://www.gov.uk/government/speeches/summary-of-updates-to-international-travel-october-2021

15. Coronavirus (COVID-19): Omicron in Scotland - evidence paper - gov.scot [Internet]. [cited 2022 Oct 6]. Available from: https://www.gov.scot/publications/omicron-scotland-evidence-paper/

16. Willett BJ, Grove J, MacLean OA, Wilkie C, De Lorenzo G, Furnon W, et al. SARS-CoV-2 Omicron is an immune escape variant with an altered cell entry pathway. Nat Microbiol. 2022 Aug;7(8):1161–1179.

17. 6 African countries added to red list to protect public health as UK designates new variant under investigation - GOV.UK [Internet]. [cited 2023 Jan 6]. Available from: https://www.gov.uk/government/news/six-african-countries-added-to-red-list-to-protect-public-health-as-uk-designates-new-variant-under-investigation

18. Coronavirus (COVID-19): international travel - gov.scot [Internet]. [cited 2022 Oct 6]. Available from: https://www.gov.scot/publications/coronavirus-covid-19-international-travel-quarantine/

19. Precautionary and temporary measures introduced to improve COVID surveillance from China - GOV.UK [Internet]. [cited 2023 Jan 5]. Available from: https://www.gov.uk/government/news/precautionary-and-temporary-measures-introduced-to-improve-covid-surveillance-from-china

20. Grépin KA, Ho T-L, Liu Z, Marion S, Piper J, Worsnop CZ, et al. Evidence of the effectiveness of travel-related measures during the early phase of the COVID-19 pandemic: a rapid systematic review. BMJ Glob Health. 2021 Mar;6(3).

21. Burns J, Movsisyan A, Stratil JM, Coenen M, Emmert-Fees KMF, Geffert K, et al. Travel-related control measures to contain the COVID-19 pandemic: a rapid review (Review). Cochrane Database Syst Rev. 2020;(9).

22. UK Civil Aviation Authority. Data and analysis | Civil Aviation Authority [Internet]. [cited 2023 Jan 5]. Available from: https://www.caa.co.uk/Data-and-analysis/

23. Managing cross-border travel during the COVID-19 pandemic - National Audit Office (NAO) Report [Internet]. [cited 2022 May 3]. Available from: https://www.nao.org.uk/report/managing-cross-border-travel-during-the-covid-19-pandemic/

24. COVID-19 reporting to include further data on reinfections - News - Public Health Scotland [Internet]. [cited 2023 Jan 10]. Available from: https://publichealthscotland.scot/news/2022/february/covid-19-reporting-to-include-further-data-on-reinfections/

25. Nicholls SM, Poplawski R, Bull MJ, Underwood A, Chapman M, Abu-Dahab K, et al. CLIMB-COVID: continuous integration supporting decentralised sequencing for SARS-CoV-2 genomic surveillance. Genome Biol. 2021 Jul 1;22(1):196.

26. R Core Team. R: A language and environment for statistical computing. R Foundation for Statistical Computing, Vienna, Austria. [Internet]. 2019 [cited 2023 Jan 17]. Available from: https://www.r-project.org/

27. Mateus ALP, Otete HE, Beck CR, Dolan GP, Nguyen-Van-Tam JS. Effectiveness of travel restrictions in the rapid containment of human influenza: a systematic review. Bull World Health Organ. 2014 Dec 1;92(12):868–880D.

28. Errett NA, Sauer LM, Rutkow L. An integrative review of the limited evidence on international travel bans as an emerging infectious disease disaster control measure. J Emerg Manag. 2020;18(1):7–14.

29. Gostic K, Gomez AC, Mummah RO, Kucharski AJ, Lloyd-Smith JO. Estimated effectiveness of symptom and risk screening to prevent the spread of COVID-19. Elife. 2020 Feb 24;9.

30. Quilty BJ, Clifford S, CMMID nCoV working group2, Flasche S, Eggo RM. Effectiveness of airport screening at detecting travellers infected with novel coronavirus (2019-nCoV). Euro Surveill. 2020 Feb;25(5).

31. Mandal S, Bhatnagar T, Arinaminpathy N, Agarwal A, Chowdhury A, Murhekar M, et al. Prudent public health intervention strategies to control the coronavirus disease 2019 transmission in India: A mathematical model-based approach. Indian J Med Res. 2020;151(2 & 3):190–199.

