## Supplementary Material for "Evaluating public health effects of risk-based travel policy for the COVID-19 epidemic in Scotland"

**Authors:** Isobel McLachlan^1,†^, Selene Huntley^1,†^, Kirstin Leslie^1,†^, Jennifer Bishop^1^, Christopher Redman^1^, Gonzalo Yebra^1^, Sharif Shaaban^1^, Nicolaos Christofidis^1^, Samantha Lycett^2^, Matthew T.G. Holden^1,4^, David L. Robertson^1,3^, Alison Smith-Palmer^1,*^, Joseph Hughes^1,3^, Sema Nickbakhsh^1,5,*^

**Affiliations**

^1^Public Health Scotland, Meridian Court, 5 Cadogan Street, Glasgow G2 6QE, United Kingdom

^2^Roslin Institute, Easter Bush Campus, University of Edinburgh, Edinburgh, EH25 9RG, United Kingdom

^3^MRC-University of Glasgow Centre for Virus Research, 464 Bearsden Road, Glasgow, G61 1QH, United Kingdom

^4^School of Medicine, University of St Andrews, North Haugh, St Andrews, KY16 9TF, United Kingdom

^5^School of Biodiversity, One Health & Veterinary Medicine, Garscube Campus, University of Glasgow, Glasgow, G61 1QH, United Kingdom

^†^Shared first authors

*Corresponding authors: Dr. Sema Nickbakhsh; Dr. Alison Smith-Palmer

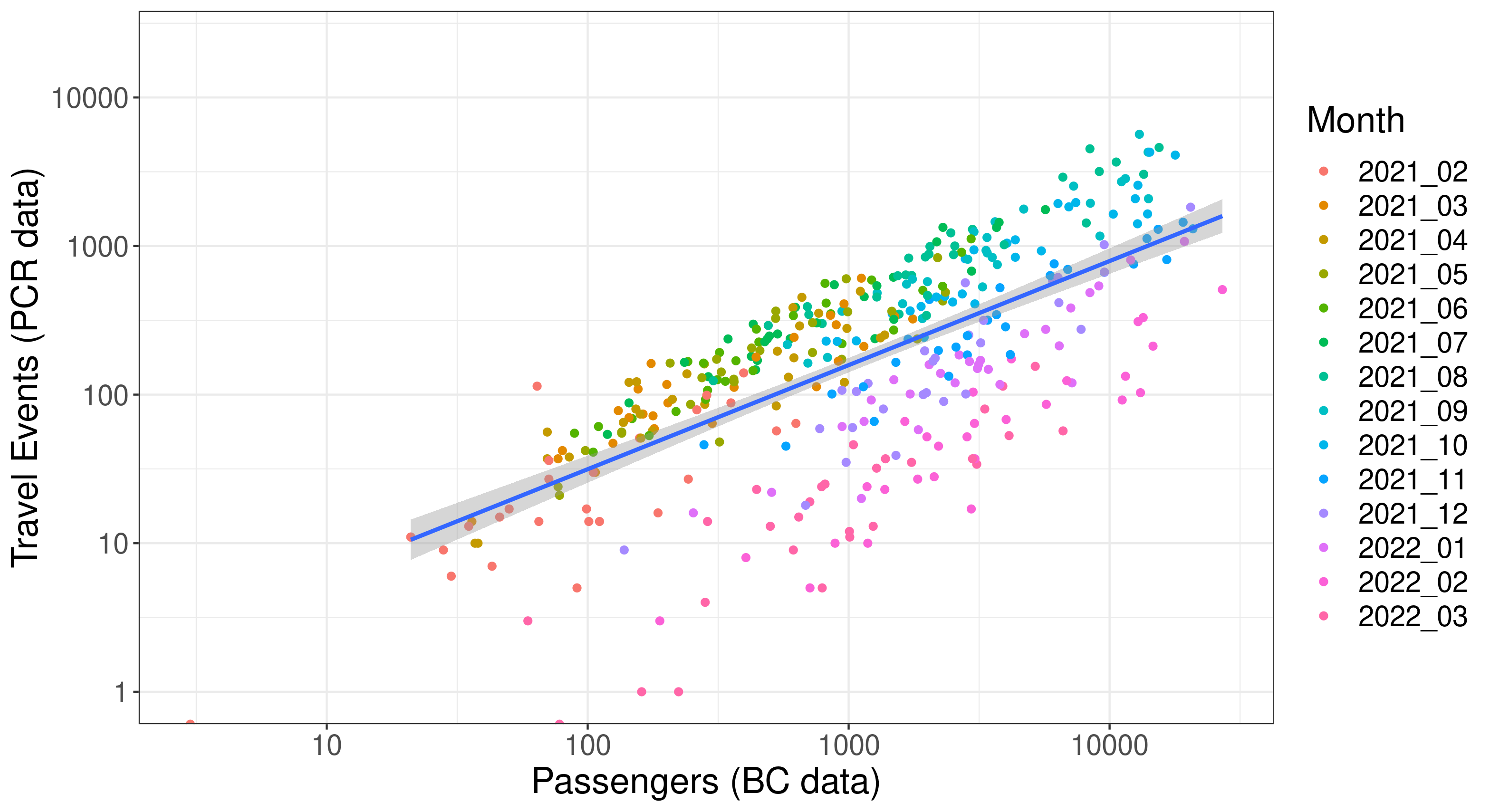

**Figure S1. Assessing correlation between data sources informing on international travel frequency.** Weekly numbers of passengers arriving into Scotland (based on Border Control Passenger Locator Forms) and international travel events (based on COVID-19 PCR-test surveillance data).

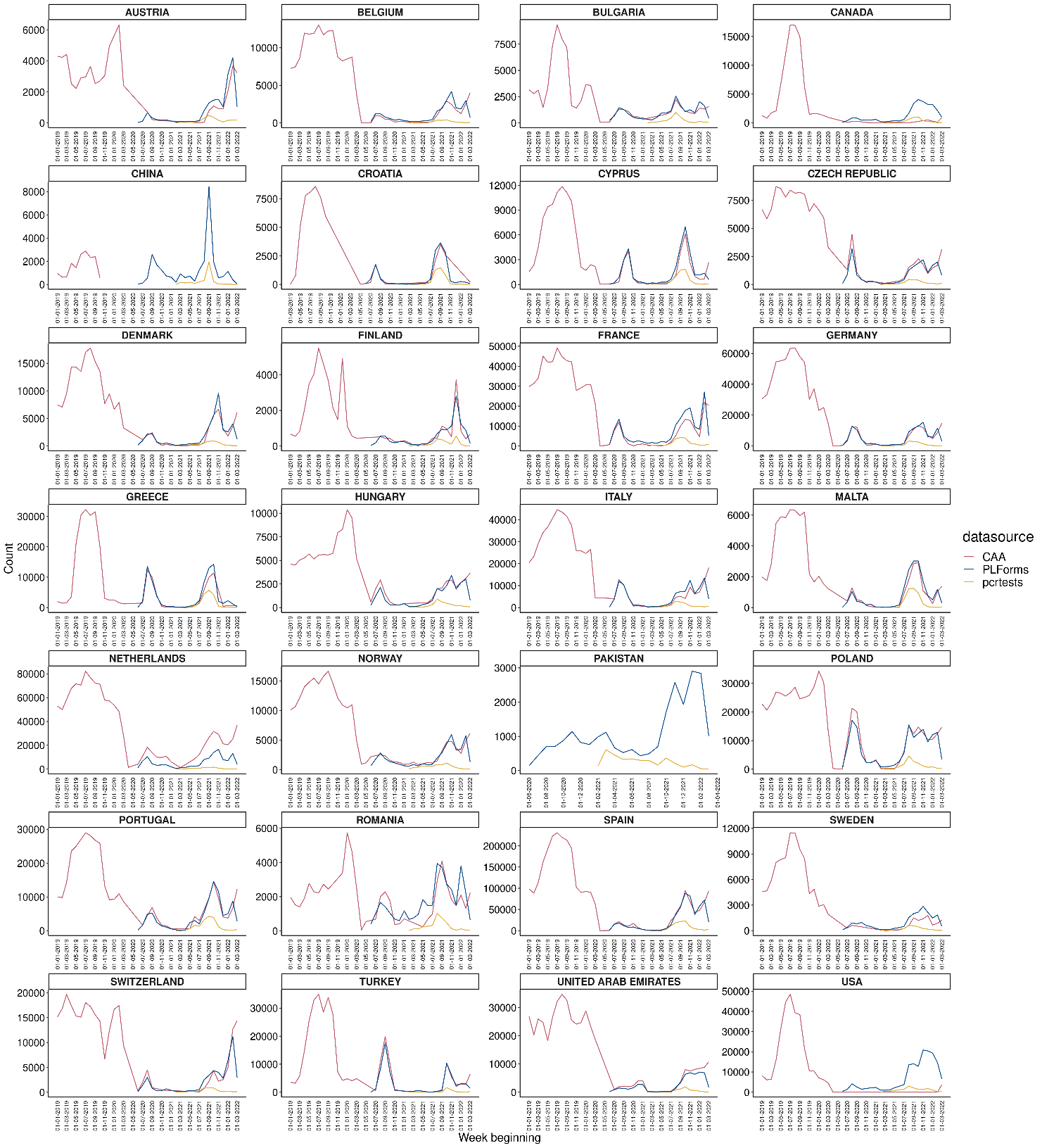

**Figure S2**. **Comparing longitudinal trends in weekly frequency of international travel by Scottish residents across data sources.** January 2019 to March 2022. BC = Border Control (Passenger Locator Forms); CAA = Civil Aviation Authority; PCR test = SARS-CoV-2 tested individuals with a recent international travel event.

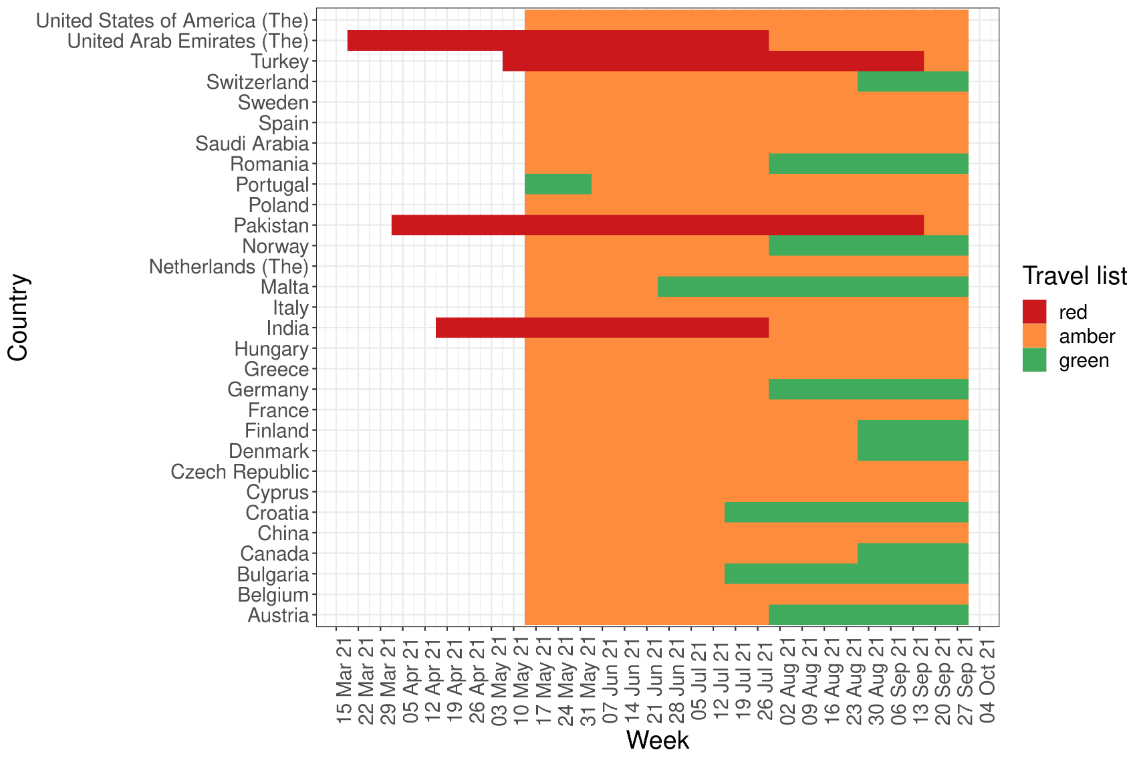

(a)

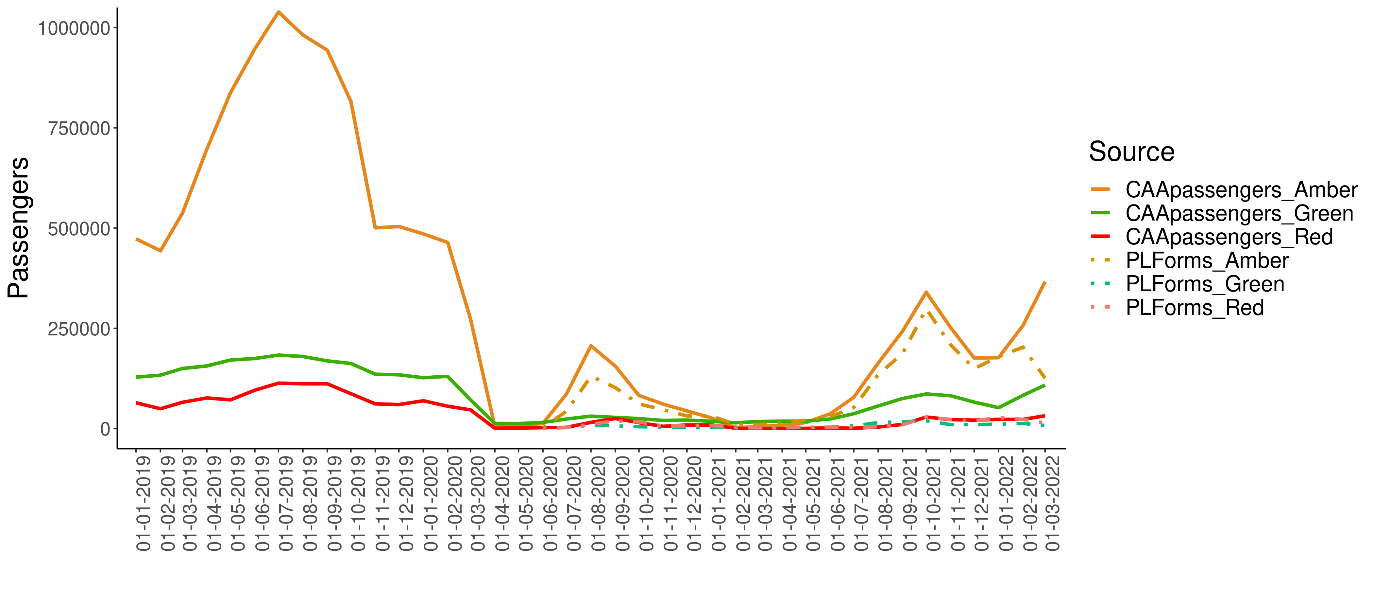

(b)

**Figure S3. Assessing the impact of the traffic light system on travel frequency in Scotland.** (a) Summary of Red-Amber-Green list designations in each week of the traffic light period shown for the top 30 most frequently visited countries (as determined based on COVID-19 PCR-tested Scottish residents). (b) Weekly numbers of passengers into Scotland retrospectively applying Red-Amber-Green (RAG) group classifications from the traffic light period to the travel destinations in Civil Aviation Authority and Passenger Locator Form datasets, to compare periods spanning pre-and-post introduction of the traffic light system.

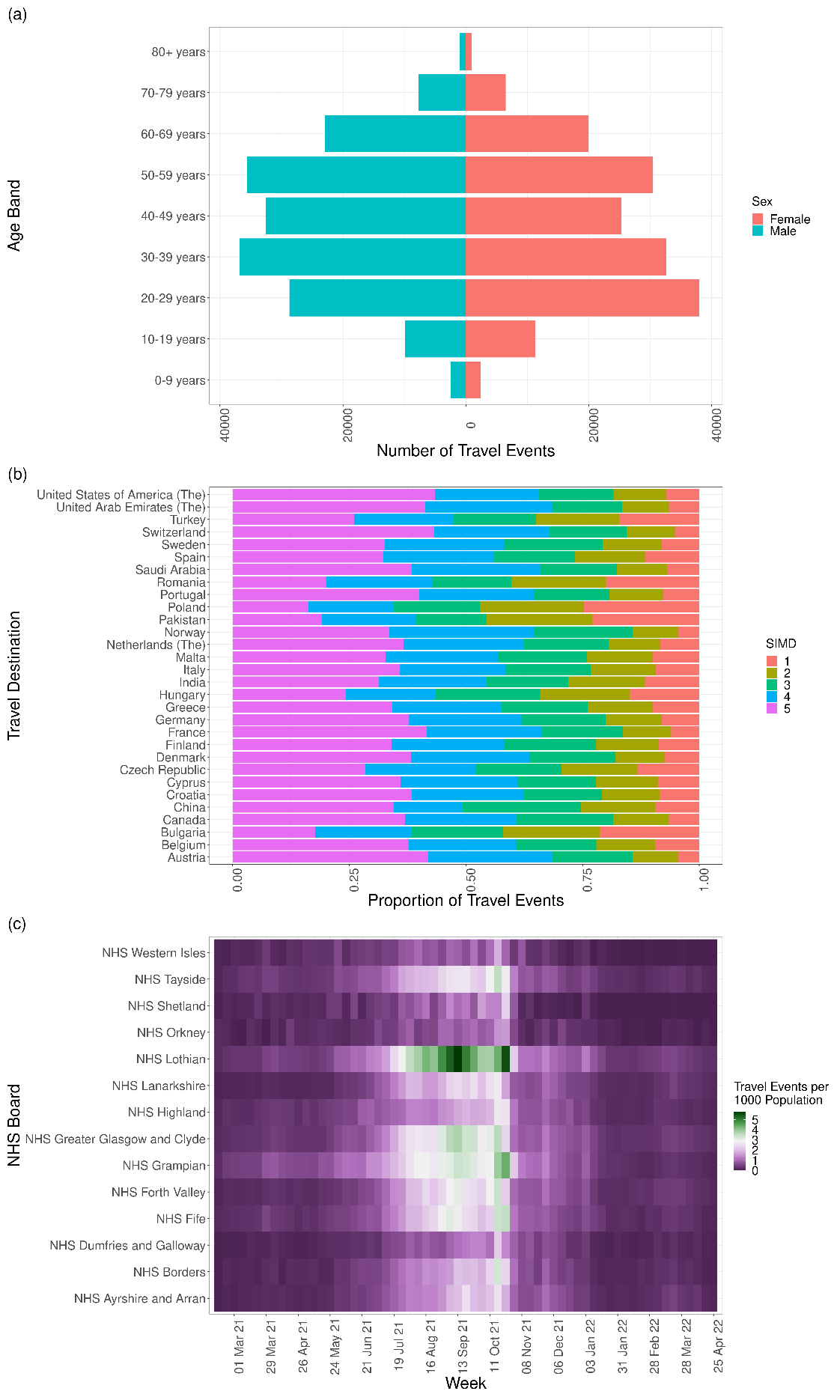

**Figure S4. Demographic and geographic distributions of Scottish residents with an international travel even within 14-days of requesting a COVID-19 PCR-test, w/c 15^th^ February 2021 to w/c 24^th^ April 2022.** Distributions are shown by (a) age-sex, (b) Scottish Multiple Index of Deprivation; SIMD (an increasing score reflects decreasing deprivation), and (c) by regional health board of residence.

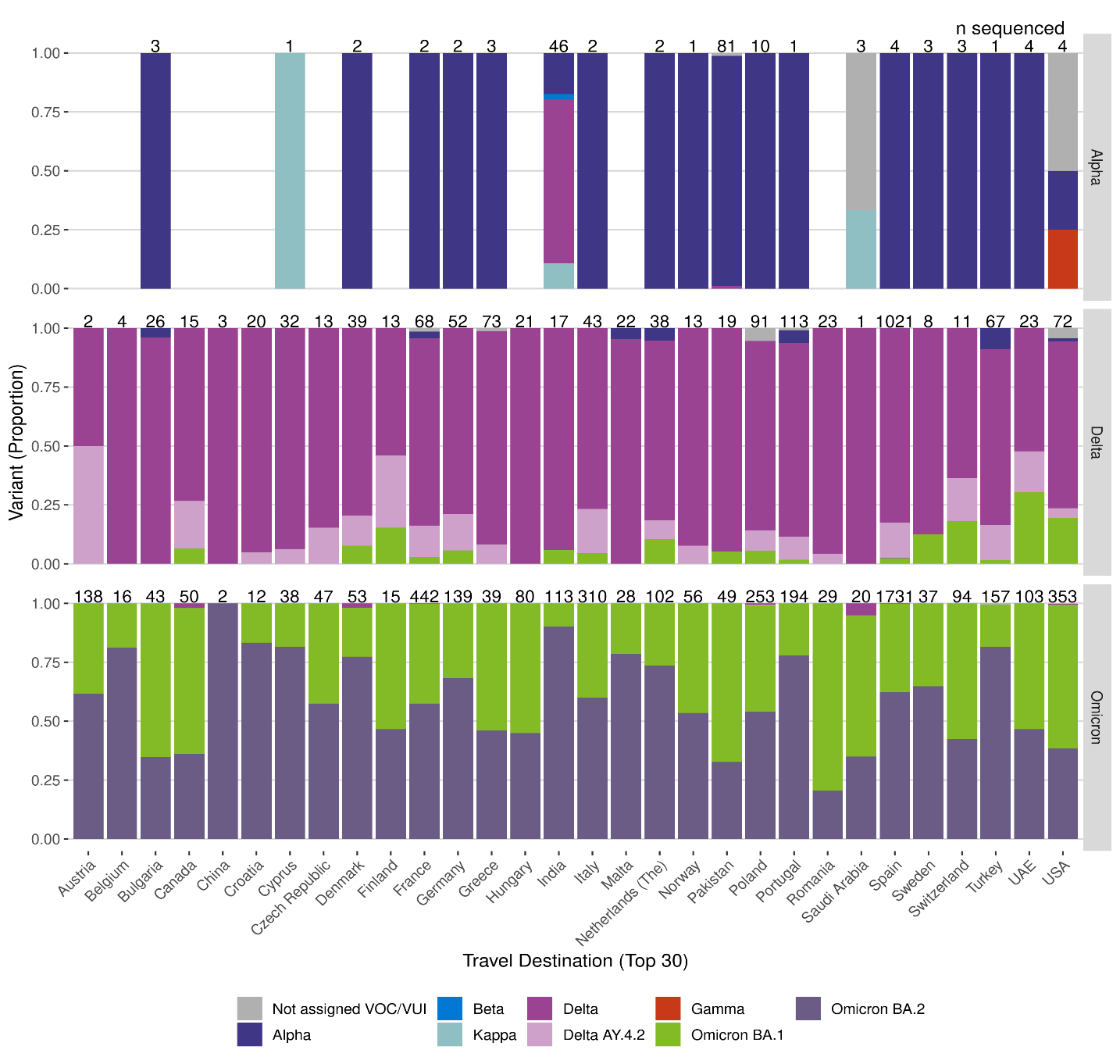

**Figure S5. Proportion of PCR-confirmed SARS-CoV-2 cases who had undergone whole genome sequencing and classified infected with a SARS-CoV-2 Variant of Concern.** Data are shown from travellers to top-30 destinations during Alpha, Delta, and Omicron eras.

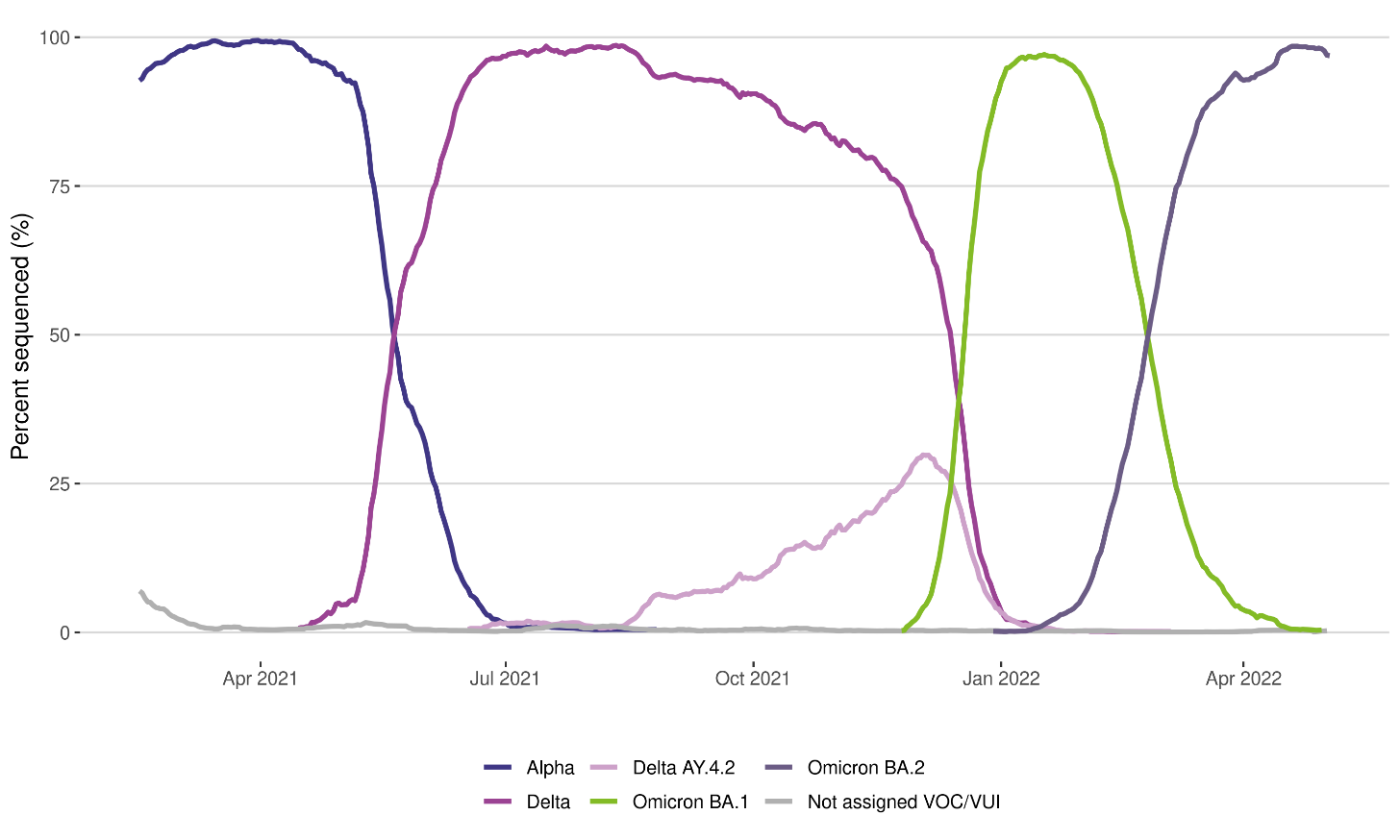

**Figure S6. Epidemic curve of dominant SARS-CoV-2 Variants of Concern identified by whole genome sequencing in the Scottish population during the study period (15^th^ February 2021 to 3^rd^ May 2022).**
